# Survival Prediction Landscape: An In-Depth Systematic Literature Review on Activities, Methods, Tools, Diseases, and Databases

**DOI:** 10.1101/2024.01.05.24300889

**Authors:** Ahtisham Fazeel Abbasi, Muhammad Nabeel Asim, Sheraz Ahmed, Sebastian Vollmer, Andreas Dengel

## Abstract

Survival prediction integrates patient-specific molecular information and clinical signatures to forecast the anticipated time of an event, such as recurrence, death, or disease progression. Survival prediction proves valuable in guiding treatment decisions, optimizing resource allocation, and interventions of precision medicine. The wide range of diseases, the existence of various variants within the same disease, and the reliance on available data necessitate disease-specific computational survival predictors. The widespread adoption of artificial intelligence (AI) methods in crafting survival predictors has undoubtedly revolutionized this field. However, the ever-increasing demand for more sophisticated and effective prediction models necessitates the continued creation of innovative advancements. To catalyze these advancements, the need of the hour is to bring existing survival predictors knowledge and insights into a centralized platform. The paper in hand thoroughly examines 22 existing review studies and provides a concise overview of their scope and limitations. Focusing on a comprehensive set of 74 most recent survival predictors across 44 diverse diseases, it delves into insights of diverse types of methods that are used in the development of disease-specific predictors. This exhaustive analysis encompasses the utilized data modalities along with a detailed analysis of subsets of clinical features, feature engineering methods, and the specific statistical, machine or deep learning approaches that have been employed. It also provides insights about survival prediction data sources, open-source predictors, and survival prediction frameworks.

## Introduction

According to World Health Organization (WHO), around ten thousand diseases have been discovered and each disease has unique symptoms, characteristics, and implications on human health^1^. Millions of people died from such diseases in the span of years 2000 to 2019, while cancers, cardiovascular, and infectious diseases persisted as the leading causes of mortality^2,3^. Extensive research on the intersection of life and technology has yielded a wide range of therapies and medications for various well-known diseases. However, the core idea behind traditional therapies and medications is based on the “one-size-fits-all”^4^. In this paradigm, a single drug is supposed to effectively treat a medical condition across a variety of patient cohorts i.e., children, old and young populations^4,5^. In-depth exploration and understanding of living organisms’ inherent biological processes reveal that high variability in genetics and drug responses make one-size-fits-all medication ineffective^4,5^.

The groundbreaking discoveries of the factors contributing to the limited effectiveness of generalized medications marked the inception of the era of precision medicine^6,7^. Precision medicine offers customization in tailored medical treatments based on an individual’s unique genetic makeup, and optimization in drug selection and dosage based on the individual’s lifestyle, and environmental factors^8^. Precision medicine’s adoption and effectiveness have been significantly enhanced by the accurate, cost-effective, and large-scale analysis of molecular information obtained through next-generation sequencing^9^.

In the realm of precision medicine, survival prediction plays a pivotal role in tailoring medical treatments to individual needs^10,11^. Survival prediction categorizes patients into distinct risk groups that enhance the efficiency of resource allocation for the patients who are likely to gain the most benefit from specific treatments^10,11^. It also enables counseling of patients and their families by predicting the expected course of the disease and potential challenges^10^. In addition to medical treatments, survival prediction offers multiple advantages in research, particularly in the area of biomarker discovery and disease understanding^12,13^. Survival prediction models provide useful information about the correlation between different features and clinical outcomes. This correlation information enables the identification of novel biomarkers associated with disease prognosis^12^. Moreover, researchers leverage survival prediction to unravel disease heterogeneity which helps to identify distinct subtypes with different survival profiles^14^. This knowledge not only aids in the stratification of homogeneous patients in clinical trials but also validates therapeutic targets by assessing their relevance in predicting patient outcomes^15^. Furthermore, it enables the longitudinal monitoring of disease progression that helps to explore critical time points and progression patterns^16^.

To expedite advancements in survival prediction research, researchers are harnessing the capabilities of AI algorithms by utilizing extensive survival-related data from public databases such as the Cancer Genome Atlas Program (TCGA)^17^, and NCI Genomic Data Commons (GDC)^18,19–24^. In addition, the diversity and heterogeneity of diseases hinder the development of a universally applicable survival prediction pipeline^14,25^.

Following the need for disease-specific predictors, there is a marathon for the development of more accurate and powerful predictors^26–28^. Figure 1 illustrates that for the advancement of survival predictors, public databases provide a spectrum of clinical data^29,30^ and encompass 9 diverse omics data modalities, including gene expression (mRNA), micro RNA (miRNA), DNA methylation, copy number variation (CNV), long non-coding RNA (lncRNA), proteomics, metabolic, whole exome sequencing (WES) and mutation^23,26,31,32^. In each data modality, there exists an array of missing values that hinder survival predictors learning. Extensive research is being conducted to impute missing values by using different techniques such as deletion, multiple, K-nearest neighbor (KNN), and median imputation^33–35^. In addition, various normalization methods are also being used to normalize feature space such as quantile, variance threshold, and rank normalizations^36^.

**Figure 1.**
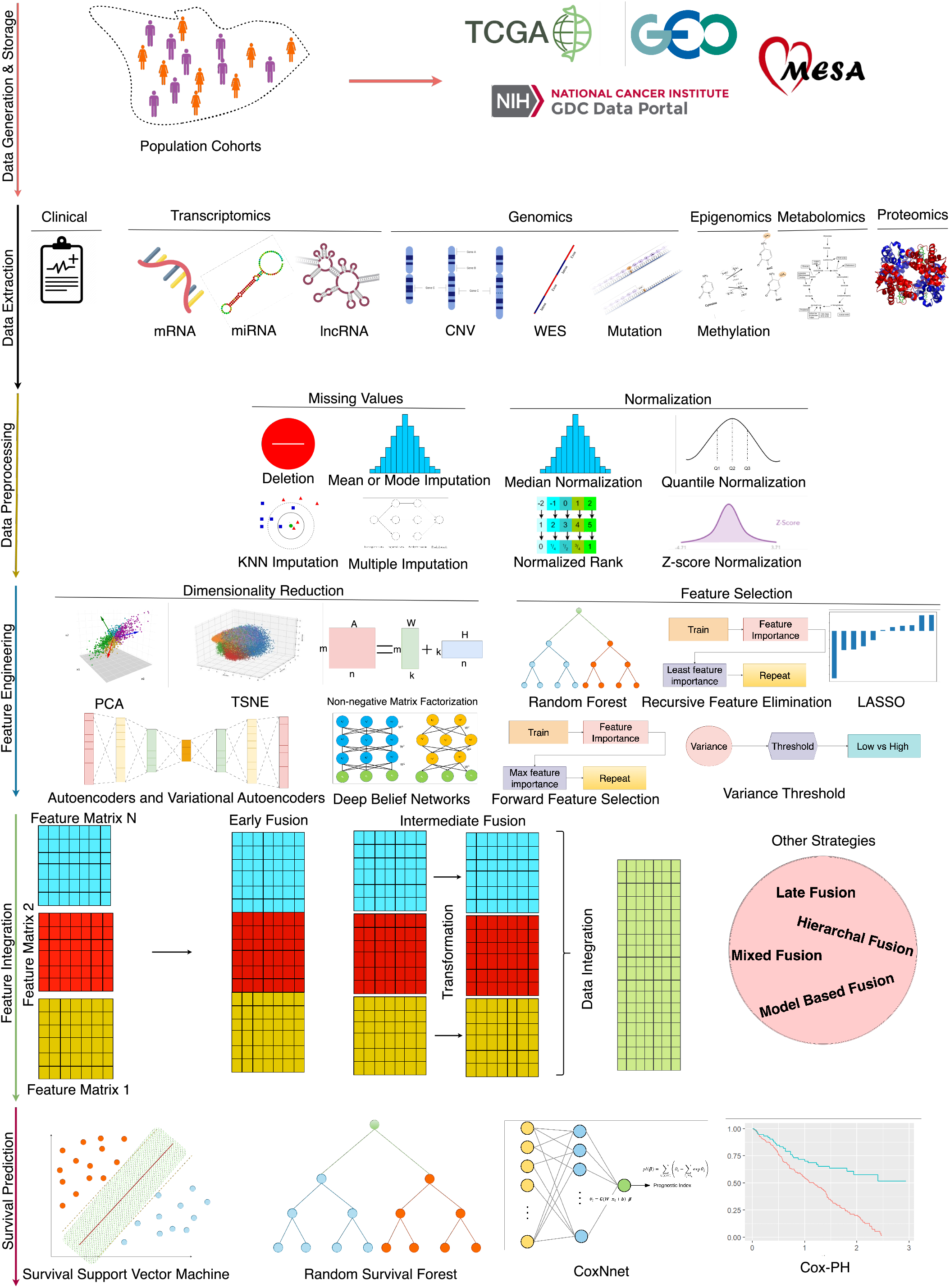
An end-to-end survival prediction pipeline.

In the development of survival prediction pipelines, researchers are trying to unlock the potential of various data modalities by assessing predictor performance with individual modalities and combinations of multiple data modalities across diverse types of diseases. When data from different modalities is combined, survival predictors’ input feature space becomes very large which impedes the performance of AI approaches^37^. Researchers are trying to explore feature engineering approaches such as random forest importance (RFI), and recursive feature elimination (RFI)^38^, principal component analysis (PCA)^31,39^, non-negative matrix factorization (NMF)^40^, and autoencoders (AEs)^41–43^. Moreover, in an end-to-end survival predictive pipeline, apart from the selection of appropriate data and feature engineering strategy, designing appropriate survival prediction models is also an active area of research.

Under different aforementioned directions, the recent 3 years have witnessed around 74 different survival predictors for different diseases. To further accelerate and expedite the development of more powerful predictors, in the last 10 years, from time to time, researchers have published 22 different review articles. These articles primarily aim to summarize the latest trends and developments in data modalities, feature engineering methods, and AI models specifically related to survival prediction. However, the focus of these reviews is often constrained to either a singular disease or multiple subtypes of cancer, highlighting a limited scope within the broader landscape of survival prediction research^37,44–48^. More comprehensive details about the scope of existing review articles in terms of contributions and drawbacks are summarised in Table 1 and section. Following the need for a comprehensive review article for survival prediction, the contributions of this paper are manifold:

**Table 1.**
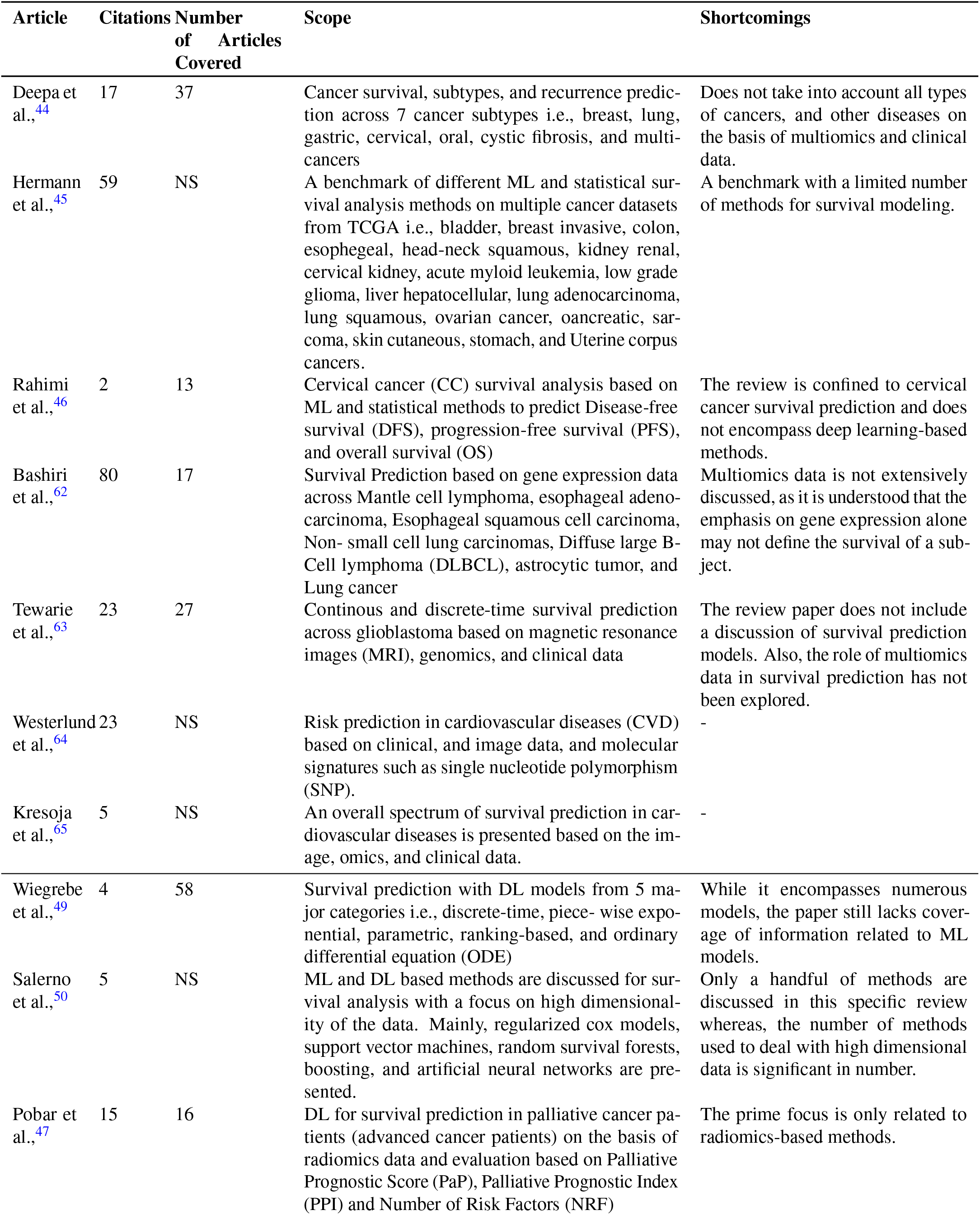
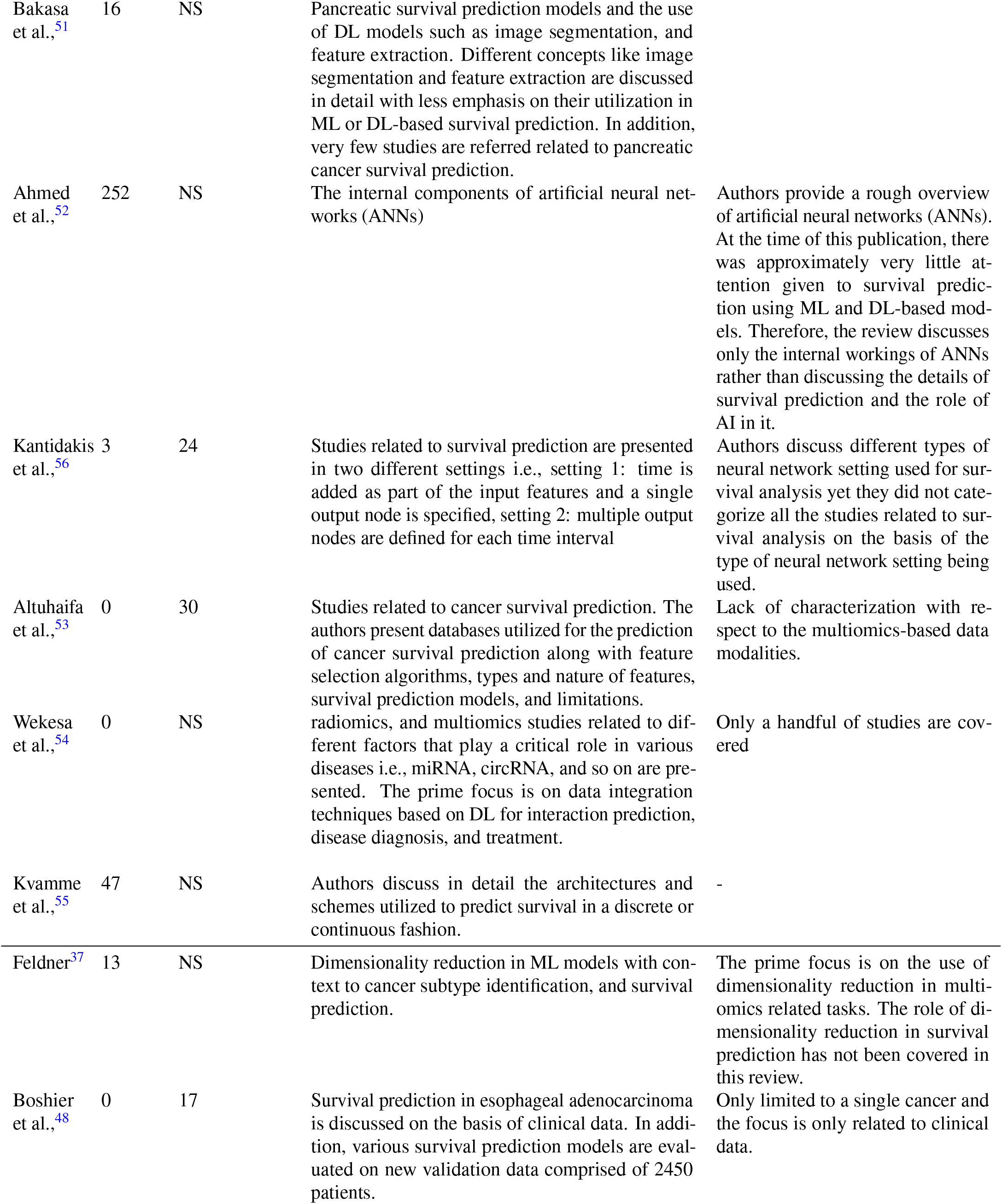
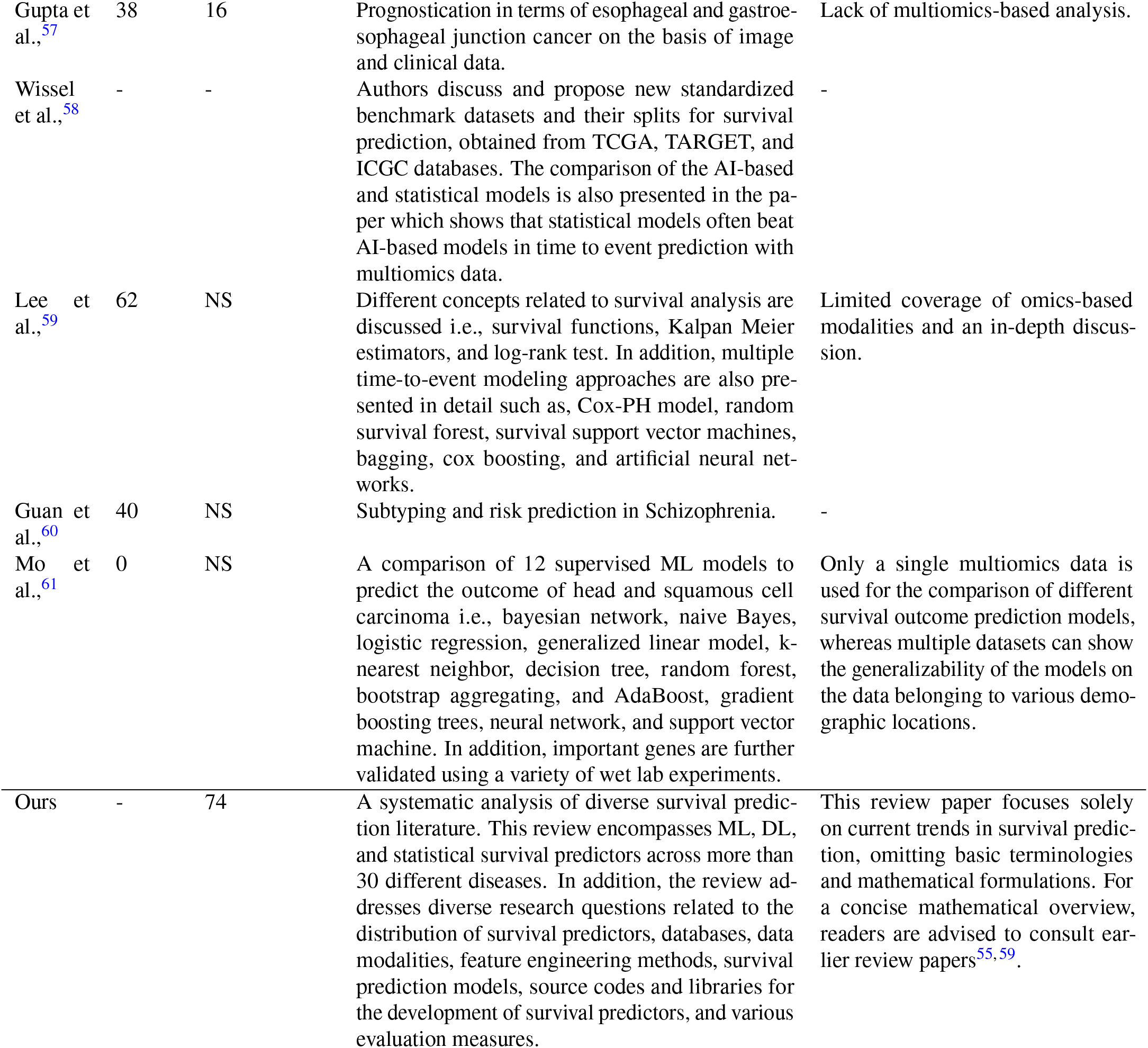
The scope and limitations of current survey papers.

- It consolidates a diverse array of 22 survival prediction review papers, bringing together their scopes and limitations under a unified umbrella. This compilation serves as a valuable resource for researchers seeking high-level insights and pertinent information in the field.
- It provides comprehensive insights into 74 survival prediction articles published between 2020 and 2023.

The objective is to delve into diverse aspects of the field, extract and furnish useful information from these articles under the following different research questions and objectives: i) What is the distribution of 74 research articles across 44 different diseases, and how does it vary among cancer subtypes and other diseases? ii) How do studies address the spectrum of survival prediction, from a broader perspective covering multiple cancer subtypes to individual subtypes? iii) What are the predominant survival endpoints used in studies, and how are studies distributed across four endpoints overall survival (OS), disease-free survival (DFS), progression-free survival (PFS), and biochemical recurrence (BC)? iv) What are the most commonly used public and private data sources in existing survival prediction studies and the types of data they encompass? v) What are the most commonly used omics data modalities and their associations with different diseases and survival endpoints? vi) Which clinical features are most commonly employed in survival prediction studies? vii) How have feature engineering techniques evolved across different data modalities, diseases, and survival endpoints in survival prediction studies? viii) Which specific statistical, machine learning (ML), and deep learning (DL) survival prediction algorithms have been applied to diverse diseases and survival endpoints? ix) Which survival prediction studies have made their source codes publicly available, and what types of methods are available in open-source survival prediction frameworks? x) What are the most commonly utilized survival prediction evaluation measures? xi) Which conferences and journals predominantly publish survival prediction studies?

## Background

Survival prediction makes use of patient-specific molecular information and clinical signatures to forecast a wide range of events at particular time intervals. The most common events include recurrence, metastasis, response, recovery, hospitalization, and progression of a disease. Some of these events represent similar contexts, i.e., metastasis and progression both contribute to the overall progression of the condition/cancer. Survival prediction events are generally categorized into 4 different survival endpoints namely, overall survival (OS), disease-free survival (DFS), progression-free survival (PFS), and biochemical recurrence (BC). Survival endpoints serve as crucial measures for assessing the outcomes of interventions, indicating the duration until specific events occur. Therefore, events are essentially the occurrences that contribute to the survival endpoints. These endpoints are critical to examine the trajectory of a particular disease.

Survival prediction is time to event approach with two distinct aspects, i.e., survival and hazard function. Survival function describes the probability that a subject survives longer than some specified time *t*. Mathematically, it is expressed as:

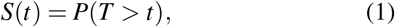

where *T* is the random variable for survival time, *t* is a specific value of interest for *T*. For instance, *S*(10) represents the probability of survival beyond 10 years without experiencing a specific event. As time passes, *S*(*t*) decreases, reflecting the reduction in the probability of surviving without the occurrence of event *E* up to time *t*.

In comparison, the hazard function illustrates the probability of an event *E* occurring at a specific time interval (Δ*t*) with a prior assumption that the event has not taken place.

The probability that the event *E* occurs within a very small time interval Δ*t* around time *t* is given by the conditional probability:

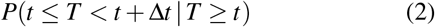

Dividing this probability by the length of the time interval (Δt) gives the rate of occurrence of the event at time *t*. The limit as the time interval (Δt) approaches zero gives the instantaneous rate of occurrence at time *t*. Mathematically, this is represented as:

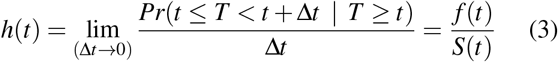

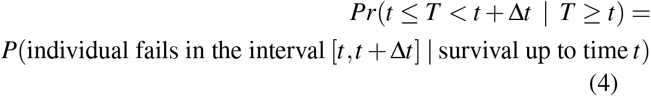

where *f* (*t*) represents the probability density function of survival time. Thus, survival function *S*(*t*) shows that the subject survives beyond a specific time point and hazard function h(t) complements this by providing a risk rate that a patient does not survive in a specific time interval conditioned on having survived thus far. Moreover, *S*(*t*) is always monotonic in nature, however *h*(*t*) is classically assumed to follow increasing Weibull, decreasing Weibull, or lognormal survival curves.

### A Look-back Into Existing Review Studies

In recent years multiple review papers have been published and the objective of each review revolves around summarising fundamental concepts in survival prediction and identifying trends in statistical, ML, and DL algorithms that have been utilized in the development of survival predictors. Table 1 illustrates a high-level overview of the existing 22 review articles in terms of their review scope and limitations. This comprehensive summary aims to assist researchers in locating specific information within relevant articles more effectively. In Table 1, a comprehensive analysis of the scope of review articles indicates that existing studies can be classified into three distinct groups. I) 9 review papers primarily focus on the application of DL algorithms in survival prediction^47,49–56^, II) 7 review papers summarise the application of ML algorithms in survival prediction^37,48,57–61^, and 6 review papers summarise survival prediction methods from three different categories namely statistical, ML, and DL methods^44–46,62–64^.

On the other hand, in the realm of disease specific survival predictors scope of existing review papers is limited. For instance, 8 papers only summarize survival predictors on single disease or subtype of cancer, i.e., cervical cancer^46^, glioblastoma^63^, esophageal adenocarcinoma^48^, esophageal and gastroesophageal junction cancer^57^, head and squamous cell carcinoma^61^, palliative cancer patients^47^, cardiovascular diseases (CVD)^64,65^, and schizophrenia^60^. Although 4 papers cover multiple subtypes of cancer but they cover only handful of 8 different subtypes such as, breast, lung, gastric, colon, esophageal, ovarian cancers and so on.

While the scope of survival prediction extends beyond multiple diseases, existing review papers fall short to summarize current trends of data modalities, feature engineering approaches and survival prediction models. For example, Deepa et al.^44^ specifically address the primary categories of data modalities used for survival prediction, namely multiomics and clinical data. However, the review does not extensively explore trends and patterns related to the 9 different omics types i.e., gene expression (mRNA), micro RNA (miRNA), methylation, copy number variation (CNV), whole exome sequencing (WES), long noncoding RNA (lncRNA), mutation, metabolic, and proteomics, or clinical features associated with distinct cancer subtypes. Similarly, Westerlund et al.^64^ do not explore the potential of multiomics data in terms of cardiovascular diseases. In addition, various review papers completely neglect to address feature engineering in survival prediction^46,47,52,56,57,62^. For instance, Feldner et al.^37^ despite their focus on dimensionality reduction, fall short in providing a comprehensive summary of current trends in feature engineering approaches with respect to diseases and data modalities. Futhermore, a small portion of these review papers cover details of few state of the art survival prediction models^49,52,56^. While current review papers summarize survival prediction pipelines partially, there is a necessity to bring diverse information into a unified platform which offers comprehensive insights into patterns and trends associated with survival prediction pipelines.

## Results

### RQ I, II, III: Survival predictors distribution analysis across diseases and survival endpoints

The primary aim of this section is to summarise the distribution of survival predictors across various diseases and survival endpoints. Predictors distribution analysis under individual diseases offers insights into the most active trends of predictors associated with specific diseases. This consolidated distribution provides a centralized platform to access valuable information about their disease of interest. Similarly, examining the distribution of articles across survival endpoints is valuable for identifying current trends in forecasting multiple events. This approach not only enhances our understanding of the current state of predictive modeling but also facilitates researchers in efficiently accessing information specific to their desired endpoints. Through this exploration, we aim to contribute to a deeper understanding of the diverse landscape of survival prediction research and its applications across various diseases and endpoints.

Table 2 illustrates disease specific predictors distribution for both cancer and other diseases respectively. In the last 3 years, 60 predictors have been designed for different cancer subtypes related survival prediction^24,104,108^ while only 14 predictors have been designed for other diseases such as cardiovascular diseases, COVID-19, and trauma^29,112,119,120^.

**Table 2.** Distribution of survival predictors across individual diseases.

| Disease Subtype | Number of Studies | References |
| --- | --- | --- |
| Nasopharyngeal Carcinoma: | 1 | 66 |
| HER2-negative metastatic breast cancer | 1 | 67 |
| Tripple negative breast cancer | 1 | 68 |
| Breast invasive carcinoma | 1 | 14 |
| Colon adenocarcinoma | 1 | 39 |
| Gastric cancer | 1 | 42 |
| Gastrointestinal cancer | 1 | 30 |
| Adult diffuse glioma | 1 | 69 |
| Invasive ductal carcinoma | 1 | 70 |
| Pancreatic cancer undergoing biliary drainage | 1 | 71 |
| Kidney renal clear cell carcinoma | 2 | 14,72 |
| Lung squamous cell carcinoma | 1 | 14 |
| Cervical Cancer | 1 | 73 |
| Neuroblastoma | 1 | 38 |
| Rectal cancer | 1 | 74 |
| Colon cancer | 3 | 75–77 |
| Liver Cancer | 1 | 41 |
| Esophageal Carcinoma | 2 | 78,79 |
| Stomach adenocarcinoma | 1 | 72 |
| Ovarian serous cystadenocarcinoma | 2 | 72,78 |
| Kidney renal clear cell carcinoma | 2 | 31,72 |
| Lower grade glioma | 1 | 80 |
| Head-and-neck squamous cell carcinoma | 1 | 72 |
| Bladder Cancer | 3 | 40,81,82 |
| Bladder urothelial carcinoma | 1 | 72 |
| Renal cell carcinoma |  | 83,84 |
| Lymphoma | 1 | 85 |
| Hepatocellular carcinoma | 3 | 43,86,87 |
| Ovarian cancer | 4 | 86,88–90 |
| Glioblastoma | 4 | 90–93 |
| Prostate Cancer | 2 | 22,94 |
| Non-small cell lung cancer | 3 | 95–97 |
| Pancreatic cancer | 3 | 26,32 |
| Breast Cancer | 6 | 23,74,90,98–100 |
| Lung adenocarcinoma | 4 | 27,101–103 |
| Pancancer | 7 | 24,28,104–108 |
| Colorectal Cancer | 1 | 109 |
| Atherosclerosis | 3 | 29,110,111 |
| Myocardial infarction | 1 | 112 |
| Stroke | 1 | 112 |
| COVID-19 | 1 | 113 |
| Cardiovascular disease | 6 | 112,114–118 |
| Liver transplant | 1 | 119 |
| Trauma | 1 | 120 |

To date, approximately more than 100 different cancer subtypes have been identified^121^. However, a deeper analysis of the last 3 years reveals that survival prediction models have been developed for only 36 distinct cancer subtypes, as outlined in Table 2. Among 36 different subtypes, most of the predictors have been designed for breast cancer, lung adenocarcinoma, ovarian cancer, and glioblastoma. On the other hand, 7 different predictors have been designed for pancancer. Notably, there is a difference between other cancer types and pancancer because under this paradigm predictors simultaneously deal with multiple cancer subtypes. For the development of pancancer based predictors, there exists public data having more than 30 distinct cancer subtypes. However, researchers are utilizing different subsets for the development of predictors. Figure 3 provides an overview of multiple survival prediction studies that encompass a range of cancer subtypes, either within a pancancer context or within the context of predicting survival for different subtypes. A total of 14 studies have taken into account multiple cancer subtypes whereas the majority of the studies have only covered only a single type of cancer subtype such as colorectal cancer^109^, lymphoma^85^, colon adenocarcinoma^39^, gastric cancer^42^ and so on.

Figures 2 and 4 illustrate predictors distribution across survival endpoints. A majority of studies 54 (79%) have OS as an endpoint of survival prediction^28,82,101,120^, whereas 7 studies have incorporated multiple survival endpoints in their analysis. Out of 7 studies, 3 studies have incorporated DFS and BC^22,26,122^. Two studies have incorporated OS, DFS, and PFS^40,108^ and 2 studies have OS, and PFS as the survival end-points^31,81^. A single study has focused on DFS only^95^, and 2 only on BC^94,117^. The rest of studies either did not explicitly specify their endpoints for survival prediction or predominantly concentrated on predicting patients’ survival outcomes without a specific focus on distinct survival endpoints.

**Figure 2.**
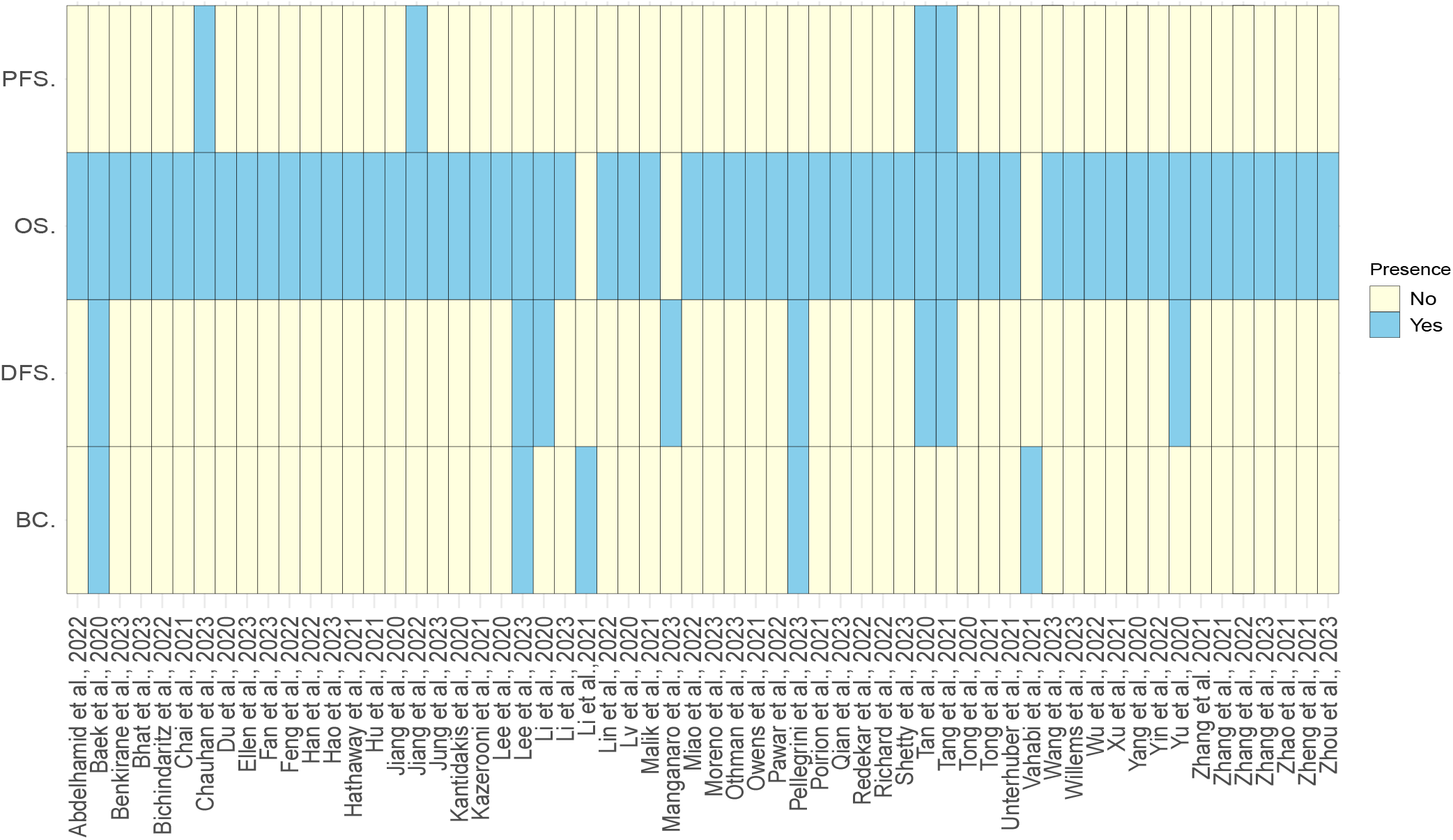
Survival endpoint distribution across diverse studies.

**Figure 3.**
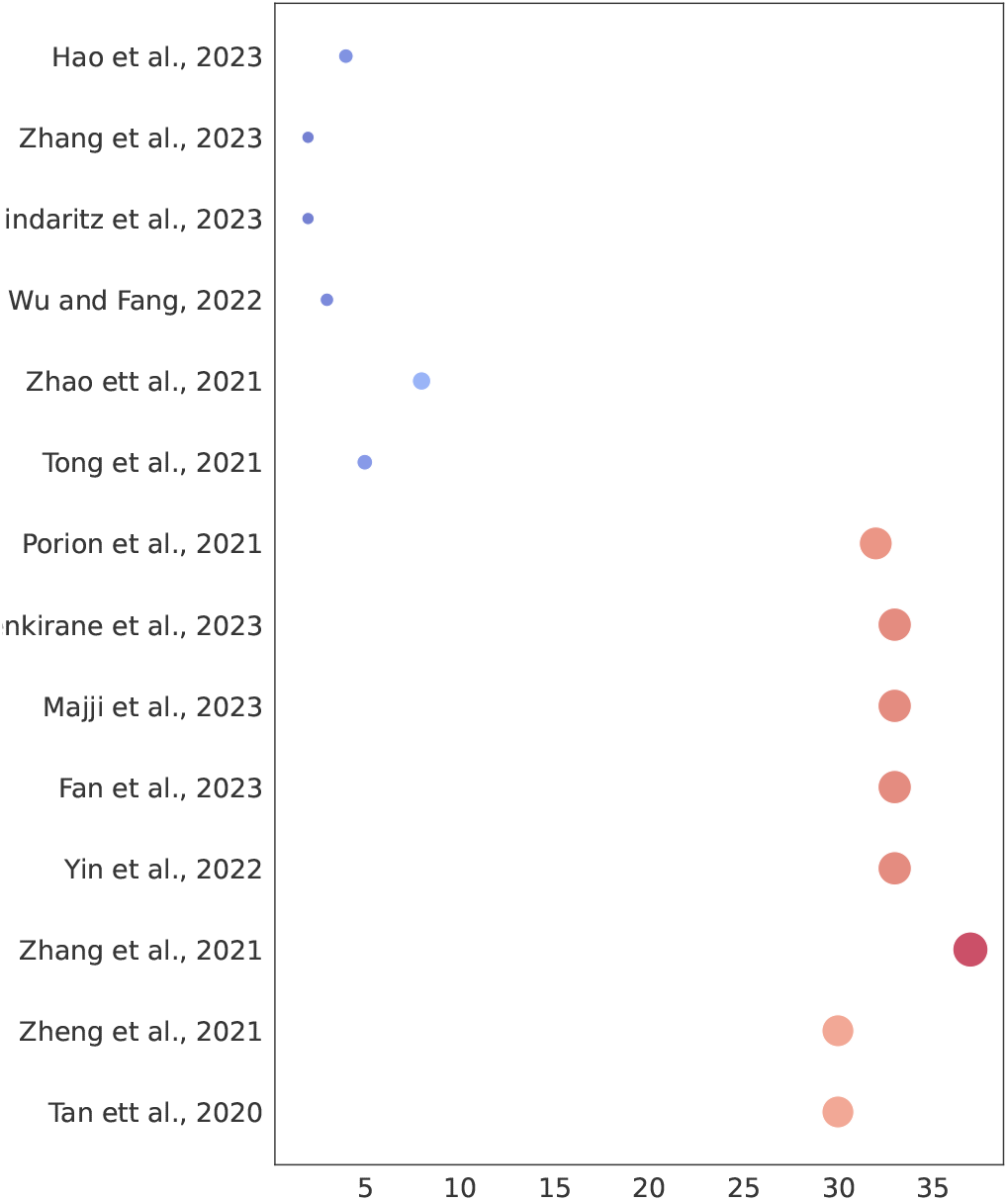
Cancer subtypes coverage based on pancancer or individual subtype settings.

**Figure 4.**
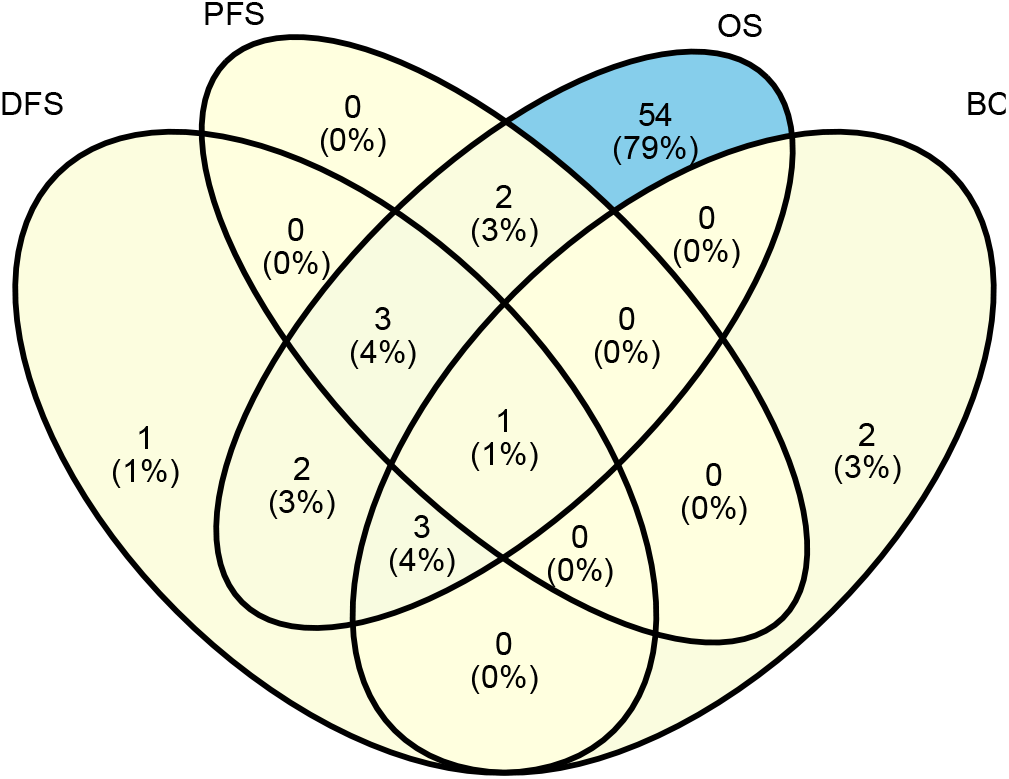
Distribution of explored survival prediction streams from existing literature. DFS: disease-free survival, PFS: progression-free survival, OS: overall survival, and BC: biochemical recurrence.

### RQ IV: Survival prediction data availability in public and private sources and opportunities for development of predictors

Survival prediction models development relies on the quality and quantity of annotated data, which is generated through extensive wet lab experiments. Experimental findings are stored in different types of databases that open new doors for the development of survival prediction applications. However, there exist multiple databases and each database encompasses particular diseases and modality specific survival data. For instance, CGGA^124^ focuses on brain tumors, and MESA^128^ contains data related to atherosclerosis. To accelerate the development of more competent survival predictors, it is essential to summarise which database contains which type of disease and what data modalities. In the highlight of research question IV, Table 3 illustrates public databases details in terms of diseases and data modalities they offer.

**Table 3.** The ample collection of survival data within diverse public databases.

| Data Source | Diseases Covered | Types of Data | URL | Description |
| --- | --- | --- | --- | --- |
| GDAC Broad Firehose<sup>123</sup> | 38 different cancer subtypes | Clinical, CNV, methylation, miRNA, mRNA, mutation, proteomics | https://gdac.broadinstitute.org/ | The Firehose platform provides processed and analyzed data from TCGA, making it accessible to researchers for further analysis and interpretation. |
| Chinese Glioma Genome Atlas (CGGA)<sup>124</sup> | Brain Tumor | Clinical, single cell RNA, mRNA, image, and microarray | http://www.cgga.org.cn/ | The Chinese Glioma Genome Atlas (CGGA) is a genomic database focused on glioma, providing comprehensive molecular characterization and clinical information to advance the understanding and treatment of glioma tumors. |
| TARGET | Pediatric cancers such as osteosarcoma, neuroblastoma, rhabdoid cancer, Wilms, acute myloid leukemia, acute lymphoblastic leukemia | Clinical, mRNA, miRNA, methylation, proteomic and CNV | https://www.cancer.gov/ccg/research/genome-sequencing/target | The TARGET (Therapeutically Applicable Research to Generate Effective Treatments) NCI (National Cancer Institute) database is dedicated to pediatric cancers, offering molecular and clinical data to facilitate research and the development of targeted therapies for pediatric cancer patients. |
| SEQC<sup>125</sup> | Neuroblastoma | Microarray, and mRNA | - | RNA-seq and microarray data to predict clinical/survival endpoints for neuroblastoma, |
| MsigDb<sup>126</sup> | - | Curated gene sets, motif gene sets, gene ontology terms, oncogenic signatures, and immunologic signatures | https://www.gsea-msigdb.org/gsea/msigdb | The Molecular Signatures Database (MSigDB) is a collection of annotated gene sets, pivotal for gene set enrichment analysis, encompassing diverse biological pathways and functions, aiding researchers in studying gene expression patterns. |
| GTEx<sup>127</sup> | 54 non-diseased tissue sites across nearly 1000 individuals | Single cell, mRNA, methylation, chip-seq, histology images | https://www.gtexportal.org/home/ | The Genotype-Tissue Expression (GTEx) project is a comprehensive research initiative that characterizes the genetic and tissue-specific gene expression patterns across a diverse set of human tissues, providing valuable insights into the relationship between genetic variation and gene regulation. |
| Kaggle | Not disease-specific | Clinical | https://www.kaggle.com/ | Kaggle is an online platform that hosts data science competitions and facilitates collaboration among data scientists, offering datasets and a community for learning and problem-solving. In addition, Kaggle is not specifically designed for omics-based datasets or studies. |
| MESA<sup>128</sup> | Heart diseases | Clinical, genetic, and imaging | https://www.mesa-nhlbi.org/default.aspx | The Multi-Ethnic Study of Atherosclerosis (MESA) MESA is a research study by the National Heart, Lung, and Blood Institute, involves 6,000+ individuals from six U.S. communities, assessed at affiliated university clinics |
| UCSC Xena<sup>129</sup> | Various cancer subtypes that are present in TCGA | Clinical and omics data modalities associated with TCGA | https://xena.ucsc.edu/ | UCSC Xena is a bioinformatics platform offering a user-friendly interface for the exploration and visualization of integrated multi-omic and clinical datasets, enabling researchers to analyze and interpret diverse biological and disease-related information collaboratively. |
| GEO<sup>130</sup> | Plethora of diseases such as cardiovascular and neurological diseases and cancers | Clinical, mRNA, miRNA, CNV, methylation, chromatin interaction, DNA modification, splicing, lncRNA, and mutation | https://www.ncbi.nlm.nih.gov/geo/ | The Gene Expression Omnibus (GEO) is a publicly accessible repository, maintained by the National Center for Biotechnology Information (NCBI), housing a diverse collection of high-throughput functional genomics datasets, enabling researchers to freely access and analyze gene expression and other omics data across a broad spectrum of biological conditions and diseases. GEO does not have a specific or dedicated portal for survival prediction datasets. |
| TCGA/GDC<sup>17</sup> | More than 40 cancer subtypes such as glioblastoma, pancreatic, bladder, breast and rectal cancers | Clinical, mRNA, miRNA, CNV, methylation, methylation, miRNA, splicing, lncRNA, and mutation | https://www.cancer.gov/ccg/research/genome-sequencing/tcga | Cancer genome atlas (TCGA) generates genomics data, while GDC is the platform that hosts and shares not only TCGA data but also other cancer genomics datasets, promoting data accessibility and collaboration in the cancer research community. |

A deeper analysis of existing survival predictors reveals that among the 74 studies 54 utilized publicly accessible data from three key databases: the Cancer Genome Atlas Program (TCGA)^17^, NCI Genomic Data Commons (GDC)^18^, and the Gene Expression Omnibus (GEO)^31, 32, 72, 73, 80, 82, 87, 90, 91, 130, 131^. Apart from public databases, there also exist private databases that have been utilized in existing survival prediction studies^66,75,81,112,113,117,118^. However, these private databases often restrict data access and may require extensive research proposals for data retrieval. Among these databases commonly used databases are Heidelberg University Hospital^30^, COMBO-01^71^, Life cohort^115^, and UNOS^119^. The reliance on private databases for survival prediction creates significant hurdles for research in several ways. Firstly, limited accessibility to such data impedes the reproducibility and verification of study findings by other researchers, hindering the validation and robustness of predictive models. Secondly, the lack of transparency and standardized access procedures for private datasets introduces challenges in benchmarking and comparing different survival prediction models. Lastly, the exclusivity of private databases may contribute to a potential bias in research outcomes, as the diversity and representativeness of the data are often compromised which impacts the generalizability of survival predictions to broader patient cohorts.

Public access to databases enables researchers to create survival benchmark datasets that fosters the development of survival prediction models. However, many researchers develop datasets without making them public which hinders transparency and the broader scientific community progress. The lack of shared data and presence of multiple datasets associated with a single disease pose a notable challenge in survival prediction. For instance, it hinders the establishment of standardized testing and benchmarking procedures for newly proposed survival prediction methods, leading to ambiguities in identifying the most advanced techniques. Moreover, recognizing the need for standardization in benchmarking survival prediction models, Wissel et al.^58^ introduced benchmark survival datasets tailored for both individual cancer subtypes and pancancer settings. These datasets are accessible at https://survboard.vercel.app/, contributing to a more uniform and transparent benchmarking framework within the survival prediction landscape. Particularly, here we emphasize the use of these datasets for benchmarking in addition to newly created datasets to have unified benchmarking for cancer-specific survival prediction models.

### RQ V, VI: Survival prediction data modalities and utilization of their combinations for disease and survival endpoints specific predictors development

Following the objective of research question V, the primary focus of this section is to investigate and provide a comprehensive summary of the various data modalities utilized in the development of diverse survival predictors. To address research question V, it describes the distribution of data modalities across predictors associated with four distinct survival endpoints, and 44 different diseases. Furthermore, in response to research question VI, it furnishes information regarding the specific clinical features utilized by various survival prediction studies.

Out of 74 different studies, data modalities details of only 68 studies are available. Within this subset, 14 studies exclusively used clinical data, 39 studies utilized multiomics data, and 15 studies investigated the combined potential of both clinical and multiomics data modalities. Moreover, based on characteristics of molecular information omics data is generally categorized into 9 different classes namely gene expression (mRNA), micro RNA (miRNA), methylation, copy number variation (CNV), whole exome sequencing (WES), long noncoding RNA (lncRNA), mutation, metabolic, and proteomics. The specifics of different predictors, in terms of variations in the combinations of clinical and various omics data modalities, are outlined in Table 4. Among 54 survival prediction studies based on multiomics, 49 studies utilized different combinations of four distinct omics types: mRNA, methylation, miRNA, and CNV^14, 26, 27, 42, 43, 69, 72, 73, 77, 82, 84, 89, 96, 97, 100, 101, 106, 108^. Only 7 studies utilized additional modalities such as whole exome sequencing (WES)^26,31^, long coding RNA (lncRNA)^31^, proteomics^22, 23, 108, 113, 115^, and mutation data^22, 23, 108, 115^.

**Table 4.**
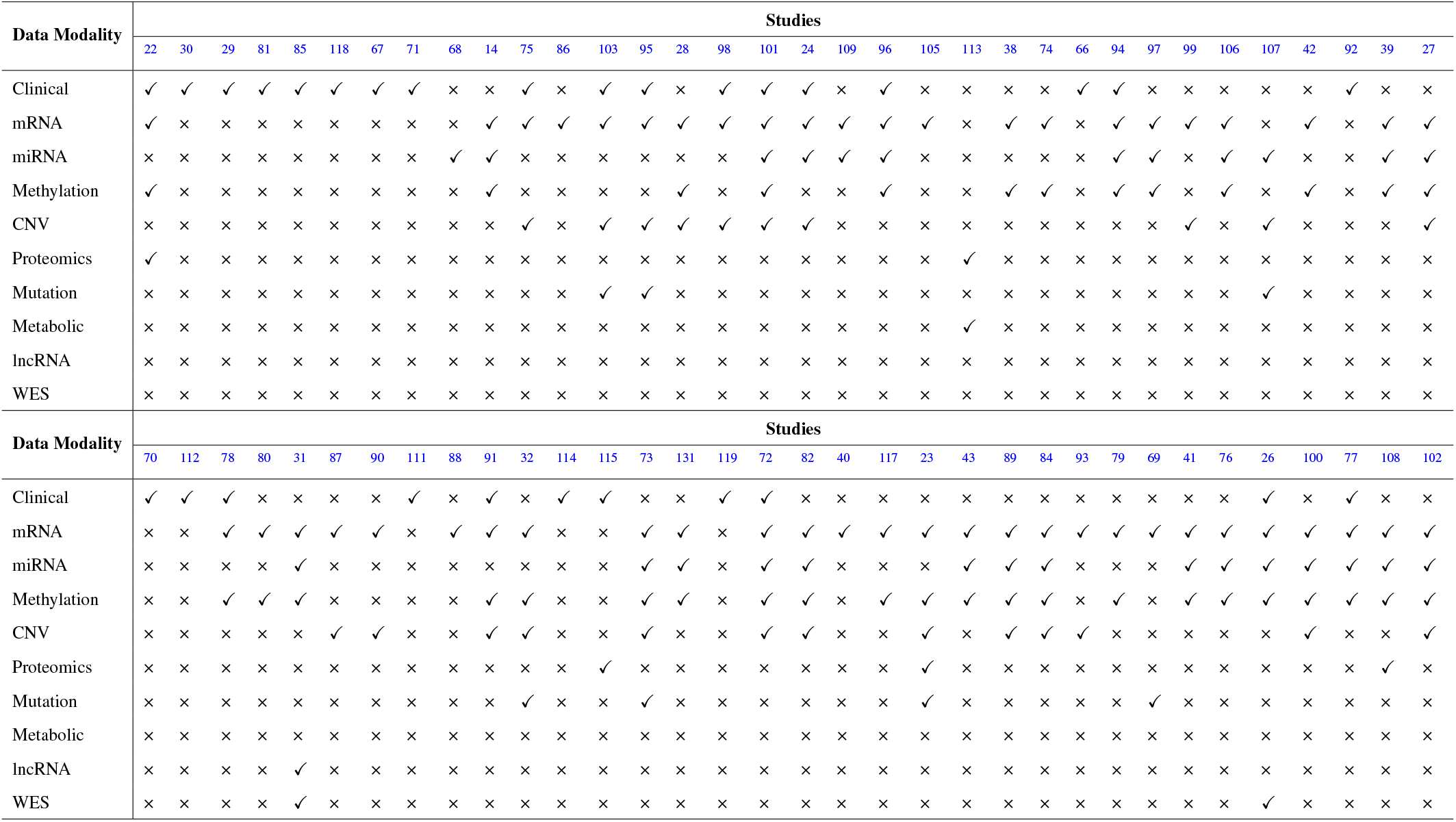
Distribution of data modalities across diverse surival prediction studies.

The choice of omics type hinges on the specific disease under investigation, as indicated by the disease-wise distribution of omics types in Figure 5. Out of 9 omics types, mRNA, CNV, miRNA, and methylation have been the most commonly utilized modalities for 33 cancer subtypes i.e., breast cancer^14, 23, 68, 74, 90, 98–100^, pancancer^24, 91, 105^–^108, 131^, colon cancer^39,74–77^, lung adenocarcinoma^27,101,102^, and ovarian cancer^72,84,88–90,103^. In addition, mutation data has been utilized for 7 cancer subtypes namely, adult diffuse glioma^69^, breast cancer^23^, cervical cancer^73^, non-small cell lung cancer^95^, ovarian cancer^103^, and pancreatic cancer^32^. Among 10 data modalities, 3 modalities namely, proteomic, lncRNA and WES have been utilized the least having limited applicability to clear renal cell cancer^31^, pancreatic cancer^26^, breast cancer^23^, localized prostate cancer^22^, and pancancer^107^. In terms of other diseases i.e., COVID-19 and heart diseases, proteomics, methylation, mRNA, metabolic, and methylation have been the only omics types utilized for survival prediction^113, 115, 117^.

**Figure 5.**
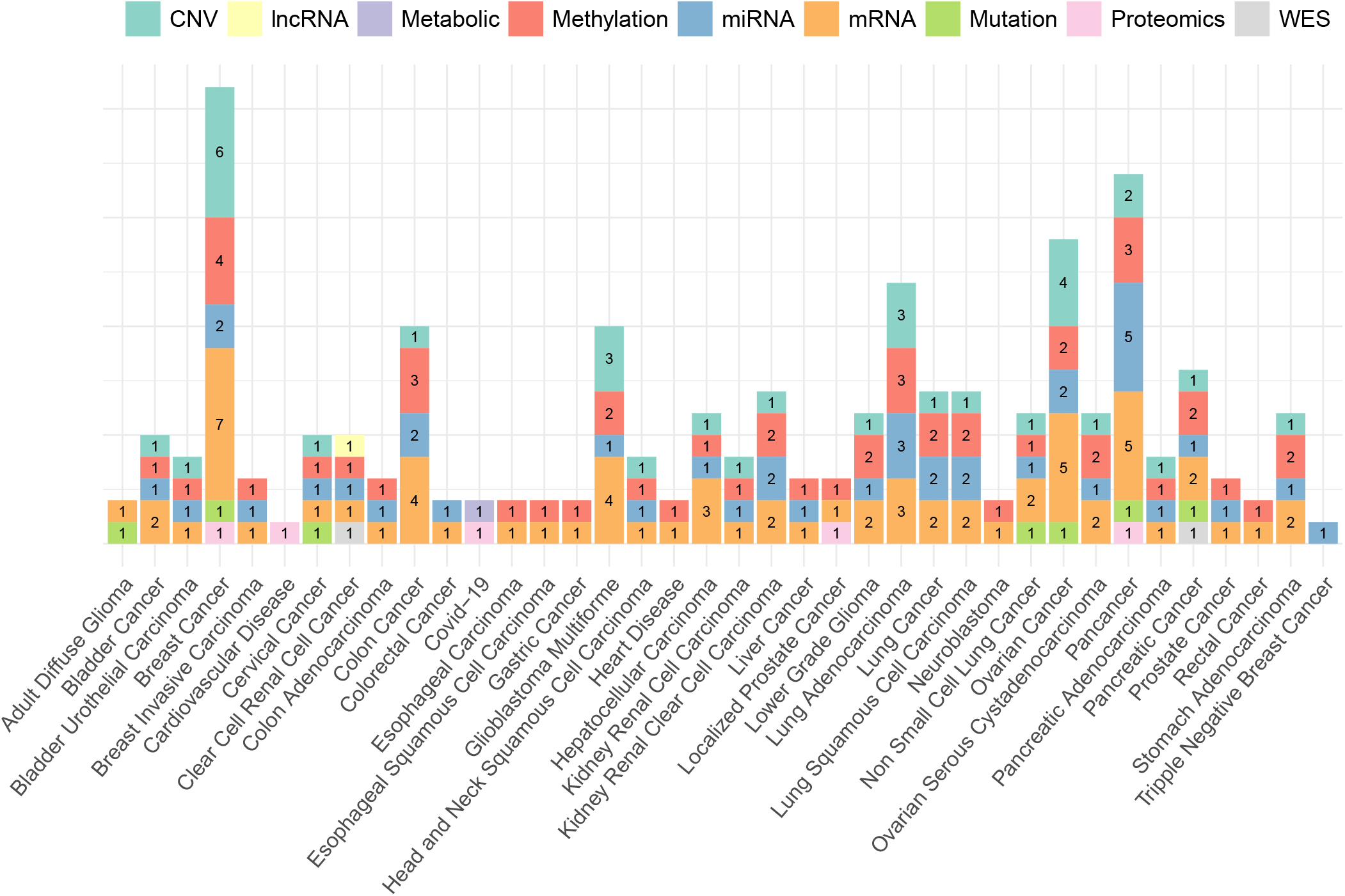
Distribution of omics data modalities across a diverse set of diseases.

The variability in omics-type selection is not solely bound to diseases but notably varies across a wide spectrum of survival endpoints. Figure 6 shows the counts of different omics types that have been utilized for different survival endpoints prediction. In the context of OS prediction, mRNA, miRNA, methylation, and CNV have been primarily utilized in more than 30 studies, with 10 studies based on proteomics, mutation, and metabolic data. However, in terms of DFS and PFS the selection of omics types appears less distinct. These endpoints have been frequently studied in conjunction with OS, predominantly utilizing mRNA, miRNA, and methylation data. This combination suggests a commonality in the predictive factors across these survival endpoints, indicating potential interconnections or shared biological processes.

**Figure 6.**
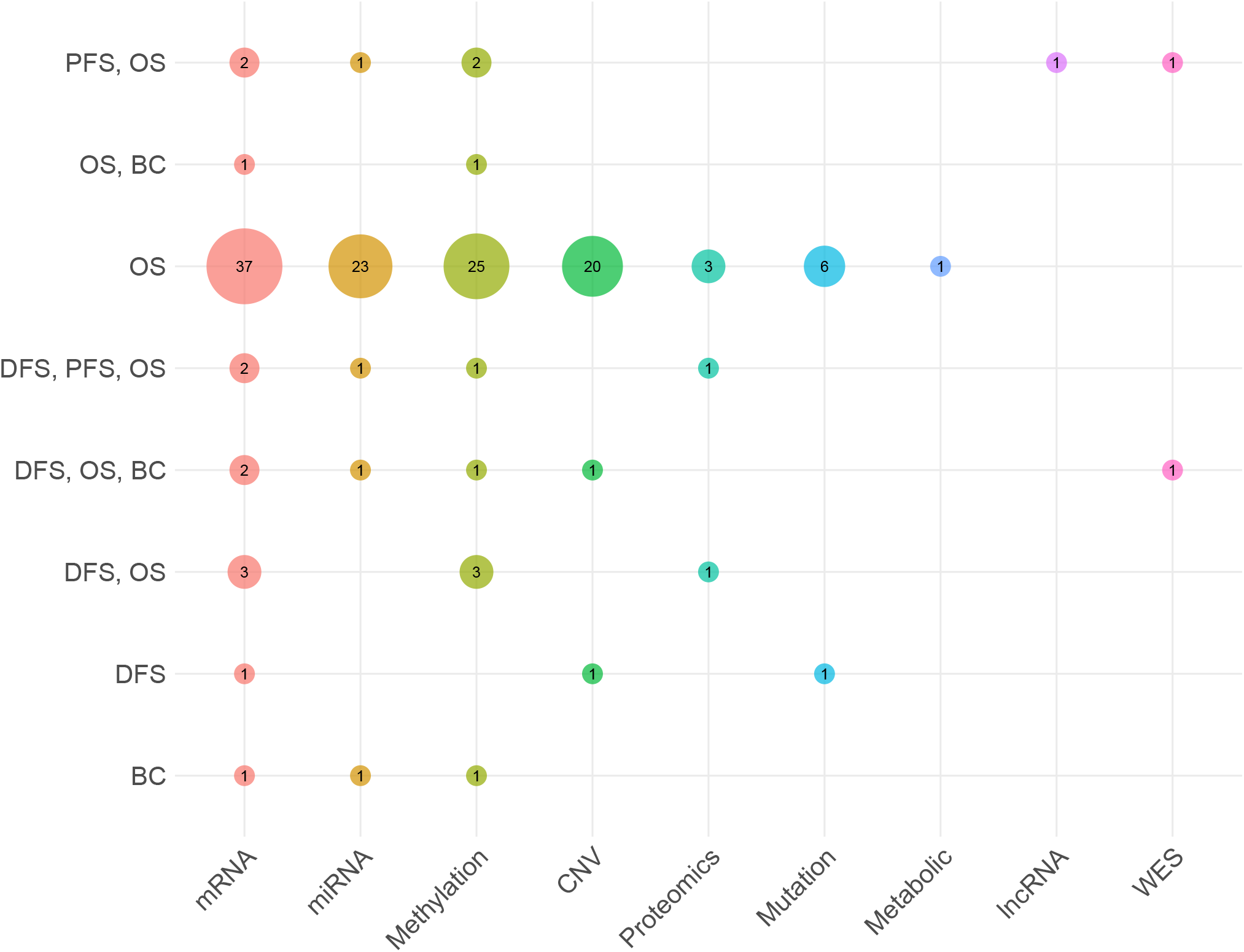
Distribution of different omics modalities with respect to survival endpoints.

Clinical data modality has been utilized in 29 different studies. However, in this modality number of features varied from study to study and it is still unclear which particular set of features is most important. To perform an in-depth analysis, which study utilized which subset of features across diverse cancer subtypes and heart diseases, a comprehensive collection of clinical features is presented in Table 5. In order to better understand and discern the trends in clinical features across diverse diseases, hereby they are placed in 7 different categories i.e., demographic features (6), disease-specific clinical markers (71), treatment-related features (17), laboratory and biomarkers (48), comorbidity and lifestyle factors (18), and other factors (15).

**Table 5.**
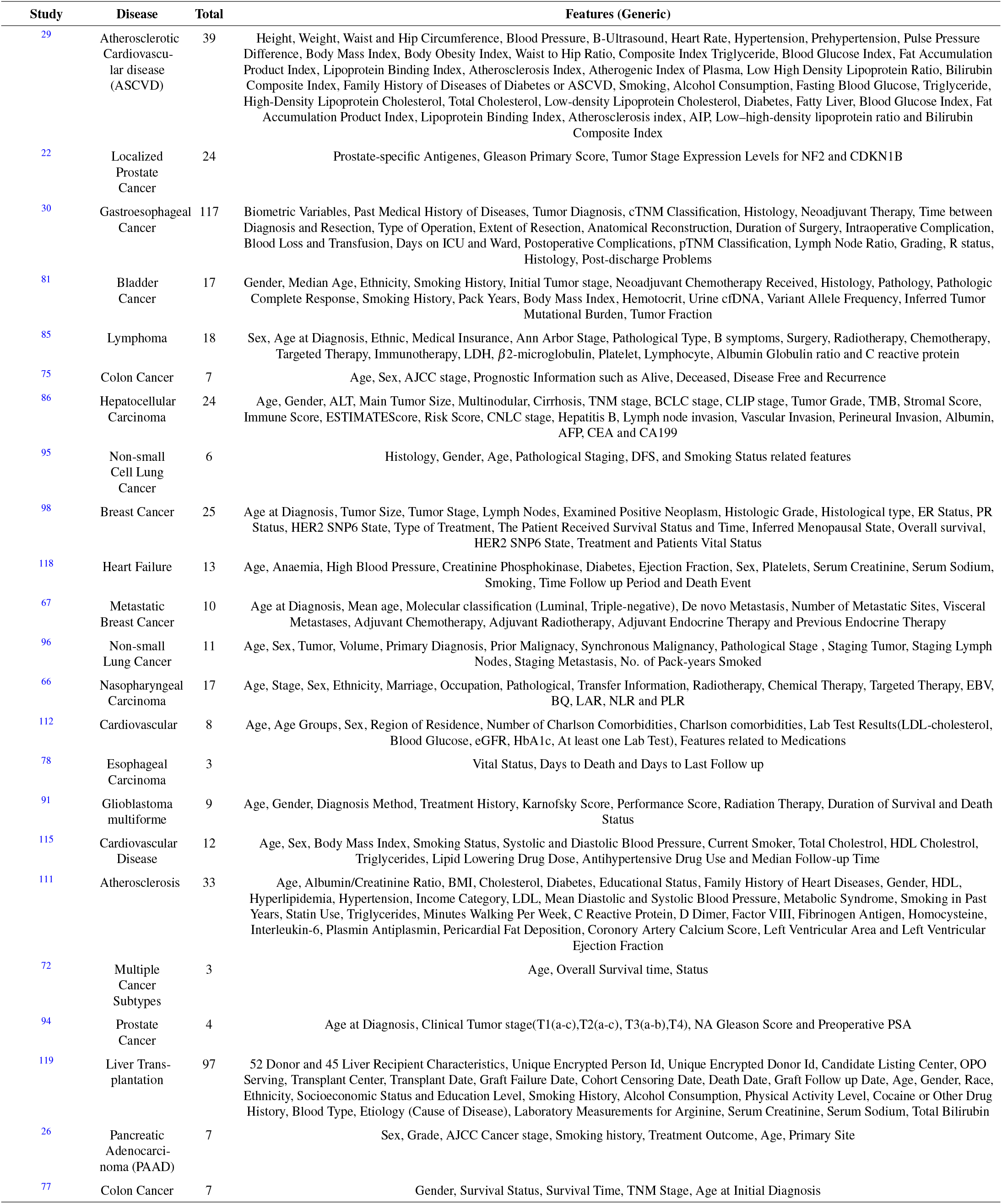
Diverse collection of clinical features utilized in various survival prediction studies.

| Study | Disease | Total | Features (Generic) |
| --- | --- | --- | --- |
| 29 | Atherosclerotic Cardiovascular disease (ASCVD) | 39 | Height, Weight, Waist and Hip Circumference, Blood Pressure, B-Ultrasound, Heart Rate, Hypertension, Prehypertension, Pulse Pressure Difference, Body Mass Index, Body Obesity Index, Waist to Hip Ratio, Composite Index Triglyceride, Blood Glucose Index, Fat Accumulation Product Index, Lipoprotein Binding Index, Atherosclerosis Index, Atherogenic Index of Plasma, Low High Density Lipoprotein Ratio, Bilirubin Composite Index, Family History of Diseases of Diabetes or ASCVD, Smoking, Alcohol Consumption, Fasting Blood Glucose, Triglyceride, High-Density Lipoprotein Cholesterol, Total Cholesterol, Low-density Lipoprotein Cholesterol, Diabetes, Fatty Liver, Blood Glucose Index, Fat Accumulation Product Index, Lipoprotein Binding Index, Atherosclerosis index, AIP, Low-high-density lipoprotein ratio and Bilirubin Composite Index |
| 22 | Localized Prostate Cancer | 24 | Prostate-specific Antigens, Gleason Primary Score, Tumor Stage Expression Levels for NF2 and CDKN1B |
| 30 | Gastroesophageal Cancer | 117 | Biometric Variables, Past Medical History of Diseases, Tumor Diagnosis, cTNM Classification, Histology, Neoadjuvant Therapy, Time between Diagnosis and Resection, Type of Operation, Extent of Resection, Anatomical Reconstruction, Duration of Surgery, Intraoperative Complication, Blood Loss and Transfusion, Days on ICU and Ward, Postoperative Complications, pTNM Classification, Lymph Node Ratio, Grading, R status, Histology, Post-discharge Problems |
| 81 | Bladder Cancer | 17 | Gender, Median Age, Ethnicity, Smoking History, Initial Tumor stage, Neoadjuvant Chemotherapy Received, Histology, Pathology, Pathologic Complete Response, Smoking History, Pack Years, Body Mass Index, Hemotocrit, Urine cfDNA, Variant Allele Frequency, Inferred Tumor Mutational Burden, Tumor Fraction |
| 85 | Lymphoma | 18 | Sex, Age at Diagnosis, Ethnic, Medical Insurance, Ann Arbor Stage, Pathological Type, B symptoms, Surgery, Radiotherapy, Chemotherapy, Targeted Therapy, Immunotherapy, LDH, $\beta_2$-microglobulin, Platelet, Lymphocyte, Albumin Globulin ratio and C reactive protein |
| 75 | Colon Cancer | 7 | Age, Sex, AJCC stage, Prognostic Information such as Alive, Deceased, Disease Free and Recurrence |
| 86 | Hepatocellular Carcinoma | 24 | Age, Gender, ALT, Main Tumor Size, Multinodular, Cirrhosis, TNM stage, BCLC stage, CLIP stage, Tumor Grade, TMB, Stromal Score, Immune Score, ESTIMATEScore, Risk Score, CNLC stage, Hepatitis B, Lymph node invasion, Vascular Invasion, Perineural Invasion, Albumin, AFP, CEA and CA199 |
| 95 | Non-small Cell Lung Cancer | 6 | Histology, Gender, Age, Pathological Staging, DFS, and Smoking Status related features |
| 98 | Breast Cancer | 25 | Age at Diagnosis, Tumor Size, Tumor Stage, Lymph Nodes, Examined Positive Neoplasm, Histologic Grade, Histological type, ER Status, PR Status, HER2 SNP6 State, Type of Treatment, The Patient Received Survival Status and Time, Inferred Menopausal State, Overall survival, HER2 SNP6 State, Treatment and Patients Vital Status |
| 118 | Heart Failure | 13 | Age, Anaemia, High Blood Pressure, Creatinine Phosphokinase, Diabetes, Ejection Fraction, Sex, Platelets, Serum Creatinine, Serum Sodium, Smoking, Time Follow up Period and Death Event |
| 67 | Metastatic Breast Cancer | 10 | Age at Diagnosis, Mean age, Molecular classification (Luminal, Triple-negative), De novo Metastasis, Number of Metastatic Sites, Visceral Metastases, Adjuvant Chemotherapy, Adjuvant Radiotherapy, Adjuvant Endocrine Therapy and Previous Endocrine Therapy |
| 96 | Non-small Lung Cancer | 11 | Age, Sex, Tumor, Volume, Primary Diagnosis, Prior Malignancy, Synchronous Malignancy, Pathological Stage, Staging Tumor, Staging Lymph Nodes, Staging Metastasis, No. of Pack-years Smoked |
| 66 | Nasopharyngeal Carcinoma | 17 | Age, Stage, Sex, Ethnicity, Marriage, Occupation, Pathological, Transfer Information, Radiotherapy, Chemical Therapy, Targeted Therapy, EBV, BQ, LAR, NLR and PLR |
| 112 | Cardiovascular | 8 | Age, Age Groups, Sex, Region of Residence, Number of Charlson Comorbidities, Charlson comorbidities, Lab Test Results(LDL-cholesterol, Blood Glucose, eGFR, HbA1c, At least one Lab Test), Features related to Medications |
| 78 | Esophageal Carcinoma | 3 | Vital Status, Days to Death and Days to Last Follow up |
| 91 | Glioblastoma multiforme | 9 | Age, Gender, Diagnosis Method, Treatment History, Karnofsky Score, Performance Score, Radiation Therapy, Duration of Survival and Death Status |
| 115 | Cardiovascular Disease | 12 | Age, Sex, Body Mass Index, Smoking Status, Systolic and Diastolic Blood Pressure, Current Smoker, Total Cholesterol, HDL Cholesterol, Triglycerides, Lipid Lowering Drug Dose, Antihypertensive Drug Use and Median Follow-up Time |
| 111 | Atherosclerosis | 33 | Age, Albumin/Creatinine Ratio, BMI, Cholesterol, Diabetes, Educational Status, Family History of Heart Diseases, Gender, HDL, Hyperlipidemia, Hypertension, Income Category, LDL, Mean Diastolic and Systolic Blood Pressure, Metabolic Syndrome, Smoking in Past Years, Statin Use, Triglycerides, Minutes Walking Per Week, C Reactive Protein, D Dimer, Factor VIII, Fibrinogen Antigen, Homocysteine, Interleukin-6, Plasmin Antiplasmin, Pericardial Fat Deposition, Coronary Artery Calcium Score, Left Ventricular Area and Left Ventricular Ejection Fraction |
| 72 | Multiple Cancer Subtypes | 3 | Age, Overall Survival time, Status |
| 94 | Prostate Cancer | 4 | Age at Diagnosis, Clinical Tumor stage(T1(a-c),T2(a-c), T3(a-b),T4), NA Gleason Score and Preoperative PSA |
| 119 | Liver Transplantation | 97 | 52 Donor and 45 Liver Recipient Characteristics, Unique Encrypted Person Id, Unique Encrypted Donor Id, Candidate Listing Center, OPO Serving, Transplant Center, Transplant Date, Graft Failure Date, Cohort Censoring Date, Death Date, Graft Follow up Date, Age, Gender, Race, Ethnicity, Socioeconomic Status and Education Level, Smoking History, Alcohol Consumption, Physical Activity Level, Cocaine or Other Drug History, Blood Type, Etiology (Cause of Disease), Laboratory Measurements for Arginine, Serum Creatinine, Serum Sodium, Total Bilirubin |
| 26 | Pancreatic Adenocarcinoma (PAAD) | 7 | Sex, Grade, AJCC Cancer stage, Smoking history, Treatment Outcome, Age, Primary Site |
| 77 | Colon Cancer | 7 | Gender, Survival Status, Survival Time, TNM Stage, Age at Initial Diagnosis |

A closer look at the clinical features across diverse diseases reveals a consistent set of fundamental demographic features i.e., age and gender which are prevalent in nearly all studies^85,86,91,111,112,115^. Beyond demographic features, disease-specific features also play critical role for disease-specific survival prediction. For instance, cancer-related studies invariably focus on tumor stage, histological type, and treatment specifics, underlining the critical role of disease-specific clinical markers in prognosis^22,75^.

Treatment-related features such as chemotherapy, radiotherapy, and immunotherapy, are particularly evident in cancer subtypes specific studies which reflect the profound influence of therapeutic interventions on survival outcomes^86,98^. Moreover, the recurrent inclusion of lifestyle and comorbidity factors ranging from smoking history and BMI to hypertension and diabetes across multiple diseases underlines their pervasive impact on prognostic modeling^101,111^. These lifestyle and comorbidity features show the complex relationship between individual health choices and their potential influence on survival outcomes.

### RQ VII: Feature engineering trends across data modalities and disease-specific survival predictors

This section addresses research question VII by investigating the application of feature engineering methods in survival prediction studies across a variety of diseases. This will help researchers to analyze and understand trends of feature engineering techniques in disease or endpoint specific survival prediction pipelines. Additionally, it delves into the trends in diverse feature engineering methods and their relevance to clinical and multiomics data modalities. This investigation aims to reveal trends and patterns in the dynamic interplay between feature engineering methods and the specific characteristics of different data modalities, and survival endpoints.

Table 6 illustrates 26 different feature engineering methods that have been utilized in diverse survival prediction studies. These methods are broadly categorized into five categories, namely supervised methods, incorporating L1 regularized Cox regression^29^, RSF algorithm^29^, Cox regression^103^, least absolute shrinkage and selection operator (lasso) regression^120^, cascaded Wx^105^, recursive feature elimination^38^, Boruta^31^, Akaike information criterion (AIC) regression^114^, variance^72^, lasso analysis^40^, multivariate regression^40^, Chi-squared^118^, mutual information^118^, and ANOVA^39,118^. Additionally, Network based methods include network based stratification (NBS)^83^, weighted correlation network analysis (WGCNA)^86^, canonical correlation analyses (CCA)^67^, patient similarity networks^38^, and neighborhood component analysis (NCA)^23^. Dimensionality reduction methods include non-negative matrix factorization (NMF)^40^, autoencoders (AEs)^28^, variational autoencoders (VAEs)^43^, principal component analysis (PCA)^39^, and dominant effect of the cancer driver genes (DEOD)^75,132^. Moreover, clustering methods comprise Kruskal-Wallis and Gaussian clustering^131^, hierarchical clustering^82^, and Guassian clustering^131^.

**Table 6.** Diverse feature engineering methods for survival prediction.

| Method Name | Studies |
| --- | --- |
| L1 regularized Cox regression | 29 |
| RSF algorithm | 29 |
| Cox regression | 29, 103 |
| Network-Based Stratification (NBS) for data integration | 83 |
| Weighted correlation network analysis (WGCNA) | 86 |
| Canonical correlation analyses (CCA) | 67 |
| Least Absolute Shrinkage and Selection Operator (lasso) regression modeling | 120 |
| Cascaded Wx | 105 |
| Recursive feature elimination | 38 |
| Patient similarity networks | 38 |
| Boruta | 31 |
| Akaike Information Criterion (AIC) regression | 114 |
| Kruskal-Wallis and Gaussian clustering | 131 |
| Variance | 72 |
| Non-negative matrix factorization (NMF) | 40 |
| Lasso analysis | 40 |
| Multivariate regression | 40 |
| PCA | 31, 39, 82, 103 |
| ANOVA | 39, 118 |
| Chi-squared | 118 |
| Mutual information | 118 |
| Hierarchical clustering | 77, 82 |
| Neighborhood component analysis (NCA) | 23 |
| DEOD | 75 |
| Variational autoencoders | 28, 89, 100, 101, 106 |
| Autoencoders | 26, 27, 31, 39, 41–43, 76, 90 |

**Table 7.** Survival analysis libraries, models, and evaluation metrics.

| Library | Language | Models | Evaluation Metrics |
| --- | --- | --- | --- |
| scikit-survival<sup>133</sup> | Python | Cox-PH, Penalized Cox-PH, RSF, Kaplan-Meier, Gradient boosting survival, Survival support vector machine | Concordance Index (C-index), Integrated Brier Score |
| Lifelines<sup>134</sup> | Python | Kaplan-MeierFitter, CoxTimeVaryingFitter, Survival regression, Discrete survival models, Piecewise exponential models | Concordance Index (C-index) |
| survival<sup>135</sup> | R | Survival regression, Cox-PH, accelerated time failure (AFT) models, Competing risk analysis, | Hazard Ratios, Log-likelihood, Akaike Information Criterion (AIC) |
| Statsmodels<sup>136</sup> | Python | PHReg, AFT models | Hazard Ratios, Log-likelihood, Akaike Information Criterion (AIC) |
| Pycox<sup>137</sup> | Python | Continuous time models such as Cox-Time, CoxCC, PCHazard and DeepSurv, Discrete time models such as Nnet-survival, probability mass function, Deep-Hit, multitask logistic regression, and BCEsurv | Concordance Index, integrated and administrative Brier Score, time dependent concordance index, negative and integrated binomial log likelihood |
| Pysurvival<sup>138</sup> | Python | CoxPH, RSF, Kaplan-Meier, Survival Support Vector Machine, multi-task logistic regression, Parametric models like exponential, Weibull, Gompertz, log logistic, and log normal | Concordance Index (C-index), Integrated Brier Score |
| flexsurv<sup>139</sup> | R | Parametric survival models (e.g., Weibull, Exponential) | Hazard Ratios, Log-likelihood, Akaike Information Criterion (AIC) |
| mlr3proba<sup>140</sup> | R | Density estimation measures, Cox-PH, flexible spline models, penalized regression, RSF, Van Belle support vector machine, gradient boostinf machine DeepSurv, DeepHit, Cox-Time | Houwelingen's $\beta$, C-index, time dependent AUC, log-loss, integrated log loss, Brier and integrated Brier score, and Schmid score |
| rstpm2<sup>141</sup> | R | Restricted Mean Survival Time (RMST), Cause-specific Hazard Models, Fine-Gray Model (Competing Risks) | IBS, Time-dependent ROC curves, Grays Test for Equality of Cumulative Incidence Functions |
| survex<sup>142</sup> | R | Local and global explanations for survival prediction models | None |

A comprehensive analysis of feature engineering methods across a range of disease-specific survival prediction studies unveils that supervised methods, such as Cox regression, L1 regularized Cox regression, and RSF algorithm, have been prevalent in diseases like ASCVD, trauma, and ovarian cancer^103,120^. On the other hand, network based methods including NBS and WGCNA, have been applied in diseases like KIRP, and hepatocellular carcinoma, which shows the significance of network structures in certain medical contexts^86^. Univariate analyses, including ANOVA, chi-squared, and univariate Cox regression, have been prevalent in diseases such as pancreatic cancer and heart failure, underscoring the significance of statistical testing in identifying relevant features^71,118^. Furthermore, dimensionality reduction methods such as PCA, and NMF have been consistently used across various diseases namely, ovarian cancer^103^, lower grade glioma^80^, colon adenocarcinoma^39^, bladder and breast cancers^40,70^. In addition, the potential of AEs, and VAEs have also been explored in diseases like glioblastoma multiforme, breast cancer, pancancer, and Lung Adenocarcinoma for feature integration and dimensionality reduction^14,28,101^.

While feature engineering methods exhibit specificity tailored to distinct diseases, their efficacy is influenced by the inherent characteristics of the utilized data. This raises the pertinent question of which particular feature engineering method proves most effective in the context of clinical and multiomics datasets. A thorough analysis of feature engineering methods and their applicability with respect to clinical and multiomics datasets reveals that methods like Cox regression, CCA, AIC, and ANOVA have been quite widely utilized in studies involving only clinical data^29,67,114,118^. These methods have been applied to clinical data for multiple reasons for instance, such methods are interpretable which is important to gain meaningful insights for healthcare professionals. Clinical data is always multifactorial, which means that multiple features of the data can lead to a specific event, and methods like ANOVA are quite efficient in analyzing such contributors. Although, such models have shown promising performance with clinical data, yet one of the drawbacks of such models is their inability to handle non-linear data which is the case in terms of multiomics data. Considering similar limitations, multiple methods such as cascaded wx^105^, RFI^38^, PSN^31^, NMF^40^, Boruta^31^, PCA^82^ variance^72^, DEOD^75^, have been utilized to handle multiomics to capture important interactions among the features and to integrate cross modalities properly. Particularly, here methods such as AEs and VAEs play a significant role and recent studies also show a growing interest in using such methods for dimensionality reduction and feature integration by such methods for multiomics and clinical datasets i.e., AEs^26, 27, 31, 39, 39, 41–43, 76, 90^, and VAEs^28, 89, 100, 106^,.

Although the selection of a feature engineering method is tied to the characteristics of the disease and the nature of the data, there is no significant evidence to suggest that it is substantially impacted by survival endpoints such as DFS, PFS, BC, and OS. This assumption arises due to the absence of a consistent pattern in feature engineering method selection across different survival endpoints. Studies, such as^95^,^39^, and^40^, demonstrate a varied use of feature engineering techniques irrespective of the specific survival endpoints (DFS, PFS, BC, or OS). This lack of uniformity implies that feature engineering method selection is driven more by the unique characteristics of the data and disease than by the nature of the survival endpoint itself.

On the basis of various trends and patterns it can be concluded that for heart diseases, univariate analyses and supervised feature engineering methods have been utilized. Conversely, in terms of cancer subtypes a mixture of dimensionality reduction methods is observed with a recent trend toward the AEs. In terms of survival datasets, the prime focus has been to use supervised methods for clinical data and multiple dimensionality reduction methods for multiomics data. Moreover, there are no conclusive remarks that feature engineering methods get affected by the survival endpoints, as the current literature also suggests a varied use of feature engineering methods regardless of the survival endpoints.

### RQ VIII: Survival Prediction Methods Insights and Distribution Across Disease Types and Survival Endpoints

In pursuit of addressing research question VIII, this section presents an overview and insights about statistical, ML, and DL algorithms that have been utilized in existing survival prediction pipelines. It succinctly examines their emerging trends across diseases and survival endpoints. This exploration aims to empower researchers in identifying gaps within disease-specific and survival endpoint-focused studies, ultimately contributing to the enhancement of survival predictive pipelines.

Table 8 provides information about 44 diseases and the corresponding survival prediction algorithms utilized in these diseases. A deeper analysis of Table 8 shows that Cox-PH and lasso Cox-PH models have been extensively utilized for disease specific survival prediction i.e., ASCVD^29,111^, bladder cancer^40,82^, colorectal cancer^74–77^, hepatocellular carcinoma^43,86,87^, ovarian cancer^88–90,103^, lung adenocarcinoma^101^, heart failure^118^, HER2-negative metastatic breast cancer^67^, pancreatic cancer^26,71^, trauma^120^, nasopharyngeal carcinoma^66^, triple-negative breast cancer^68^, lymphoma^85^, breast cancer^40,81,82^, ovarian cancer^88–90,103^, and lower-grade glioma^80^, cardiovascular disease^112,114–117^, invasive ductal carcinoma^70^, liver transplantation^119^, gastric cancer^42^, lung cancer^27^, esophageal squamous cell carcinoma^79^, glioma^69^, and liver cancer^41^. RSF has been employed in 13 studies for 6 diseases namely, ASCVD^29^, bladder cancer^82^, gastrointestinal cancer^30^, cervical cancer^73^, liver transplantation^119^, and heart failure^118^. DL model DeepSurv, has been utilized in 5 studies related to gastrointestinal cancer^30^, ASCVD^111^, NSCLC^97^. On the other hand, in the analyzed survival predictive pipelines less frequently utilized methods are i.e., survival SVM^79,95,120^, partial logistic regression^70,75^, log hazard net^75,104^, boosting^41,112^, stepCox^86^, elastic net^95^, CNN-cox^104^, DeepOmix^104^, ordinal Cox-PH^78^, DeepHit^112^, and linear multitask logistic regression (MTLR)^112^.

**Table 8.** Distribution of survival predictors across diverse diseases.

| Studies | Disease | Predictor |
| --- | --- | --- |
| 29,111 | ASCVD | Cox-PH, RSF, MTLR, DeepSurv neural network, lasso Cox-PH |
| 14,83 | KIRP | GeneNet, ANNs |
| 22,94 | Prostate cancer | Coherent Voting Network (CVN), Best Linear Unbiased Prediction (BLUP) |
| 30 | Gastrointestinal cancer | DeepSurv, MTLR, Gompertz model, RSF |
| 68 | Tripple negative breast cancer | lasso Cox-PH |
| 40,81,82 | Bladder cancer | Cox-PH, RSF, CoxNet, and transfer learning-based CoxNet |
| 85 | Lymphoma | lasso-Cox-PH |
| 74–77 | Colon cancer | Loghazard Net, partial logistic regression, Cox-PH |
| 43,86,87 | Hepatocellular carcinoma | Stepwise Cox (StepCox), SurvivalSVM, Cox-PH, CoxNet |
| 88–90,103 | Ovarian cancer | Cox-PH, Cox-Time, and DeepSurv with consensus training |
| 95–97 | NSCLC | SVM, Elastic net and Cox-PH, CNN and ANN |
| 28 | Multiple cancers | ANN |
| 23,90,98,100 | Breast cancer | CoxNet, Cox-PH, Cox-Time, and DeepSurv with consensus training, Loghazard Net, partial logistic regression |
| 101 | Lung adenocarcinoma | Cox-PH, and lasso Cox-PH |
| 24,72,104–108,131 | Pancancer | Survival neural network, CNN-Cox, Cox-PH, DeepOmix, lasso and group penalized Cox-PH, VAE based NN |
| 109 | Colorectal cancer | Lasso Cox-PH |
| 118 | Heart failure | Cox-PH, RSF |
| 67 | HER2-negative metastatic breast cancer | Cox-PH |
| 26,71 | Pancreatic cancer | Cox-PH, l2 regularized regression |
| 120 | Trauma | RF, SVM for outcome prediction |
| 66 | Nasophrngeal carcinoma | Cox-PH |
| 112,114–117 | Cardiovascular disease | Survival outcome prediction based on naive Bayes, ANNs, and SVM, Logistic regression and XGboost. Survival prediction: Cox-PH, survival XGboost, DeepHit, DeepSurv, Cox-PH, Linear MTLR, and RSF |
| 113 | COVID-19 | SVM |
| 38 | Neuroblastoma | Deep neural network (DNN) |
| 70 | Invasive ductal carcinoma | Multivariate Cox two way stepwise regression |
| 78 | Stomach, Esophageal carcinoma and Ovarian serous cystadenocarcinoma | Bidirectional LSTM, ordinal Cox model network and auxiliary loss |
| 80 | Lower grade glioma | lasso Cox-PH |
| 31 | Renal cell carcinoma | Cox-PH |
| 90–93,102 | Glioblastoma | Cox-PH, CoxNet, SVM and Cox-PH, lasso Cox-PH |
| 73 | Cervical cancer | RSF, and Cox-PH |
| 84 | Ovarian and breast cancer |  |
| 119 | Liver transplantation | RSF, Cox-PH, and partial logistic artificial neural networks (PLANN) |
| 39 | Lung adenocarcinoma |  |
| 42 | Gastric cancer | lasso, univariate and multi-variate Cox-PH |
| 27 | lung cancer | Cox-nnet |
| 79 | esophageal squamous cell carcinoma | Support vector machine, K-means clustering |
| 69 | Glioma | Cox regression |
| 41 | Liver cancer | XGBoost for subtype classification, and Cox-PH for survival prediction |

Furthermore, supplementary Table S3 provides details about predictors distribution with respect to survival endpoints. A detailed analysis reveals, out of 74 predictors, 31, 8, 1, and 6 models have been utilized for OS, DFS, PFS, and BC survival endpoints respectively. Unlike disease-specific predictors, here a mixture of methods is utilized and no particular trend exists. To provide high-level overview of multiple methods that have been utilized in all four survival endpoints we have provided a graphical representation of methods in Figure 7.

It can be seen in Figure 7, diverse types of methods that have been utilized in survival predictive pipelines can be categorized into three different categories i.e., statistical, ML, and DL. Statistical methods are broadly classified into three different categories i.e., parametric, semi-parametric, and non-parametric models. Parametric methods make assumptions about the survival time distribution^122,143^. Parametric methods include exponential, Weibull, log-normal, Weibull, gamma models, and so on^143,144^. Comparatively, semiparametric methods make no assumptions about the shape of the baseline hazard function (non-parametric). Rather, these methods assume a specific functional form for the effect of covariates (parametric)^145^. In comparison, non-parametric methods do not take into account assumptions about the underlying distribution of survival times and the shape of the hazard function. These methods include Kalpan-Meier, Nelson-Aalen, Breslow, Gehan-Eilcoxon, and life table methods^146^. Statistical methods have certain disadvantages with multiomics based survival prediction^59^. For instance, statistical models assume linear relationships among variables and fail to capture complex and non-linear data patterns^147^. These methods perform poorly on high dimensional data where the number of features is larger than the number of samples. This specific gap is filled by the emergence of AI based models. Various ML models are utilized for survival analysis such as random survival forest^148^, and boosting-based methods^149^. Belle et al., Shivasmy et al., and Khan et al.,^150–152^ proposed ranking and regression-based survival SVM for survival prediction while handling right censored data. Particularly, survival SVM is used in three ways for survival prediction i.e., ranking, regression, and combined. Ishwaran et al.,^148^ proposed RSF where log-rank test is utilized for the splitting as compared to the Gini impurity of the classical random forest models.

**Figure 7.**
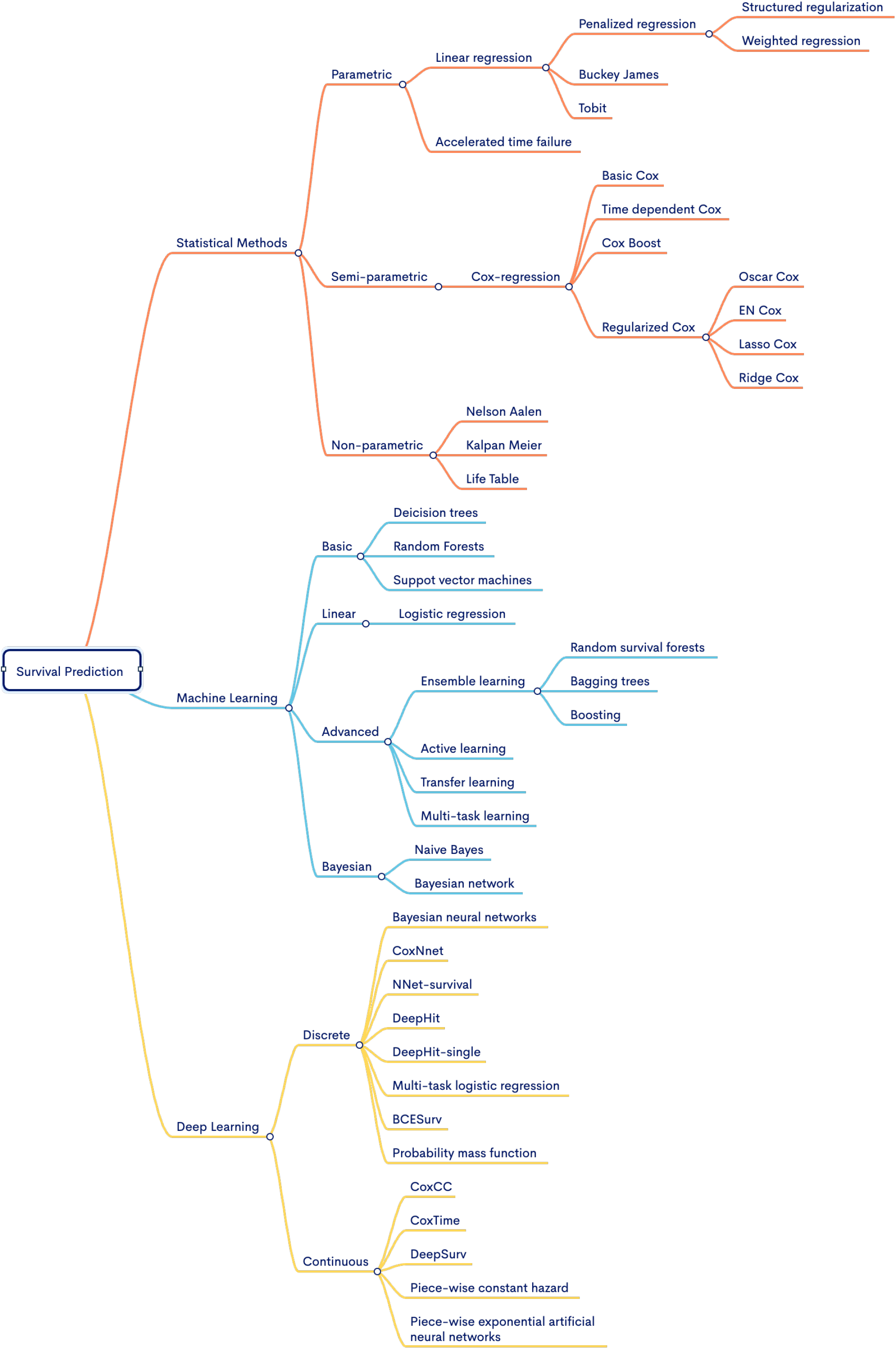
Hierarchal illustration of survival prediction methods under three different categories.

DL methods are utilized in two ways to model survival prediction tasks i.e., continuous and discrete time. Models like CoxCC and time^137^, piecewise constant hazard or PEANN^153^, and DeepSruv^154^ are utilized for continuous survival time prediction. Whereas, Nnet-survival^155^, Nnet-survival probability mass function (PMF)^156^, DeepHit and DeepHit Single^157^, multi-task logistic regression (MTLR)^158,159^, and BCESurv^160^ are utilized to predict survival in a discrete-time setting.

### RQ IX: Open source tools and libraries potential for development of survival prediction pipelines

Following the objective research question IX, this section summarizes details of open-source libraries and source codes of existing survival predictors. This comprehensive information will facilitate researchers to build upon existing work, fostering a collaborative environment and accelerating the development of robust and effective survival prediction models.

Table 9 presents an overview of open-source survival prediction models. Among the 74 distinct survival prediction studies, only 26 have provided publicly accessible source code. Among these studies, 6 studies have utilized R^91,94,96,103,109,119^ and 20 have opted for Python^14, 24, 28, 38, 72, 74, 82, 83, 89, 90, 95, 100, 105, 111, 116, 118, 131 23, 27, 106^. A comprehensive analysis of open source codes reveals that a majority of these tools have been developed from scratch without utilizing any specific survival prediction library^14, 28, 83, 95^.

**Table 9.** Summary of open-source survival prediction methods in existing studies.

| Publication | Disease | Approach | Source Code |
| --- | --- | --- | --- |
| 83 | Kidney Papillary, Renal Cell Carcinoma (KIRP) | GeneNet | Link |
| 22 | Localized prostate cancer | Coherent Voting Network (CVN) | Link |
| 14 | GBM, KRCCC, LSCC, BIC | ANNs for binary survival class prediction | Link |
| 103 | Ovarian Cancer | DT, RF, and ANN | Link |
| 95 | Non-small Cell Lung Cancer | Two layer SVM | Link |
| 28 | Pancancer | ANN | Link |
| 24 | Pancancer | Survival neural network | Link |
| 109 | Colorectal cancer | Lasso penalized cox model | Link |
| 118 | Heart failure | Two-way survival prediction | Link |
| 96 | Non-small cell lung cancer | Elastic net and cox proportional hazard model | Link |
| 105 | Pancancer | CNN and a cox model (CNN-Cox) | Link |
| 38 | Neuroblastoma | Deep neural network (DNN) | Link |
| 74 | Breast cancer | Loghazard Net | Link |
| 90 | Glioblastoma, ovary and breast cancers | CoxNet | Link |
| 91 | Glioblastoma multiforme | Cox regression | Link |
| 111 | Atherosclerosis | Various models | Link |
| 116 | Cardiovascular disease | DeepSur, Cox-PH, RSF | Link |
| 131 | Pancancer | Cox-PH model | Link |
| 72 | 8 cancer subtypes | DeepOmix based on DNN | Link |
| 82 | Bladder cancer | Cox regression, deep cox neural network | Link |
| 89 | Ovarian cancer | Cox-PH regression | Link |
| 84 | Ovarian, lung, kidney, and pancreatic cancer | Various survival models | Link |
| 119 | Liver transplantation | RSF, Cox-PH, PLANN | Link |
| 27 | Lung adenocarcinoma | Cox-nnet | Link |
| 100 | Breast cancer | DNN and cox proportional hazard model | Link |
| 94 | Prostate cancer | BLUP | Link |
| 23 | Breast cancer | Deep neural network | Link |
| 106 | Pancancer | Deep Neural network | Link |

Approximately 10 different survival prediction packages or libraries have been developed. Each library offers a diverse set of preimplemented statistical, ML, and DL survival prediction models. For instance, Pycox^137^ primarily focuses on continuous and discrete DL survival prediction models such as CoxTime, CoxCC, MTLR, and so on. Lifelines^134^, scikit-survival^133^, and pysurvival^138^ cover a wide range of statistical and ML survival prediction models like Cox-PH, RSF, survival support vector machine, and gradient boosting survival^133, 134, 138^.

Notably, addressing the lack of interpretability or explainability in the previously discussed libraries, Spytek et al.^142^ introduced Survex. This library allows researchers to analyze the features responsible for a specific event by offering different methods for both local and global explanations of various survival prediction models.

The selection of a specific library is inherently subjective and depends on factors such as the preferred development platform, choice of survival prediction models, and the specific research question in hand. Therefore, recommendations are made based on the number of survival prediction models and evaluation measures each library offers. For Python, Lifelines^134^ and Pycox^137^ are recommended, with Lifelines^134^ providing a diverse range of statistical and ML models, while Pycox^137^ is specialized in DL models. Additionally, for R, mlr3proba^140^ is recommended, as it offers a variety of statistical and ML models for survival prediction. Ultimately, selecting a library aligned with individual research needs not only streamlines the development process but also contributes to the overall reliability of survival prediction models.

### RQ X: Strategies for assessing survival predictors: unveiling common evaluation measures

The main objective of this section is to provide a concise overview of research question X, which focuses on the commonly employed evaluation measures for survival predictive pipelines.

Table 10 shows a compilation of 18 distinct evaluation measures that have been commonly used to evaluate survival prediction pipelines. The survival prediction pipelines can be categorized into two distinct classes namely survival outcome prediction and survival prediction. Details related to these categories is provided in the background section. Out of 18 evaluation measures mentioned in Table 10, a set of 10 evaluation measures have been employed to assess the performance of survival outcome prediction models. In addition to the aforementioned measures, 8 other evaluation measures have been utilized to assess the performance of survival prediction models.

**Table 10.** A summary of evaluation measures used in survival prediction and survival outcome prediction pipelines.

| Task | Evaluation Measure | Count | References | Advantages |
| --- | --- | --- | --- | --- |
| Survival Prediction | C-index | 43 | 22,29,83 | Robust, measures discriminatory power. Less sensitive to censoring compared to other metrics. |
|  | BS | 7 | 111,116,119 | Measures accuracy of predicted survival probabilities. |
|  | IBS | 5 | 30,98,116 | Considers entire survival time distribution. |
|  | Log Rank P-value | 9 | 28,31,68,91 | Tests differences in survival experiences. |
|  | DCA | 1 | 66 | Accounts for clinical consequences. |
|  | Kappa | 3 | 22,83,107 | Measures agreement beyond chance. |
|  | TD-ROC | 2 | 67,68 | Time-dependent evaluation of ROC. |
|  | AUC-pval | 1 | 22 | Evaluates AUC significance. |
|  | Odds ratio | 1 | 22 | Measures association between groups. |
|  | Likelihood Ratio | 1 | 77 | Helps in understanding the odds of a predicted event occurring compared to the odds of it not occurring. |
| Survival Outcome Prediction | AUC-ROC | 21 | 14,22,29 | Provides a comprehensive view of the model's performance across various threshold values. |
|  | Accuracy | 12 | 14,75 | Simple and easy to understand, providing an overall measure of correct predictions. |
|  | Precision | 6 | 14,67,81,98 | Useful when the cost of false positives is high, as it focuses on the accuracy of positive predictions. |
|  | Recall | 6 | 14,67,81,98 | Emphasizes the ability of the model to capture all positive instances, important for sensitive scenarios. |
|  | MCC | 1 | 98 |  |
|  | F1-Score | 2 | 38,114 | Harmonizes precision and recall, making it useful when there is a trade-off between false positives and false negatives. |
|  | PPV | 1 | 81,114 | Focuses on the proportion of true positives among positive predictions, providing insights into prediction accuracy. |
|  | NPV | 1 | 81,114 | Focuses on the proportion of true negatives among negative predictions, providing insights into prediction accuracy. |

In survival prediction category based evaluation measures the objective is to capture two distinct characteristics namely, discrimination and calibration. Specifically, calibration refers to how well the predicted probabilities of survival align with the actual observed survival rates over time. Under this paradigm most widely used evaluation measures are BS, IBS, TD-ROC, and DCA. Discrimination paradigm based evaluation measures capture differentiation between individuals with different survival times. Under this paradigm most widely used measures are C-index, AUC-ROC, and likelihood ratio. On the other hand objective of survival outcome prediction evaluation measures is to assess diverse characteristics of a model i.e., efficacy of the model, overall accurate predictions, biasness towards type I or type II errors. Specifically, accuracy and F1 score are used to measure overall accurate predictions, precision, and recall examine the model’s biasness with respect to type I and type II errors. Additionally, MCC provides a comprehensive assessment, taking into account overall accurate predictions, and errors. In addition, AUC-ROC assesses the predictive potential of a model by analyzing the true positive rate (TPR) and true negative rate (TNR) at different thresholds.

### RQ XI: Publisher and journal-wise distribution of research papers

This section addresses research question XI by presenting the distribution of survival prediction literature across diverse journals and publishers. Overall, this analysis not only enables researchers to strategically position their work but also offers opportunities for interdisciplinary collaboration, promoting a more interconnected and dynamic research landscape within the domain of survival prediction.

In Figure 8 and 9, the distribution of survival prediction literature is presented based on journals and publishers. The studies have been published in 16 different publishers, including but not limited to Springer, Elsevier, Oxford Press, and BioMed Central. Notably, around 30 out of 74 survival prediction studies have been disseminated through Springer, and BioMed Central. Furthermore, Elsevier has contributed to the field by publishing 10 relevant papers in recent years. Particularly, these studies have been published in more than 50 different conferences/journals, which shows the diversity of the survival prediction landscape.

**Figure 8.**
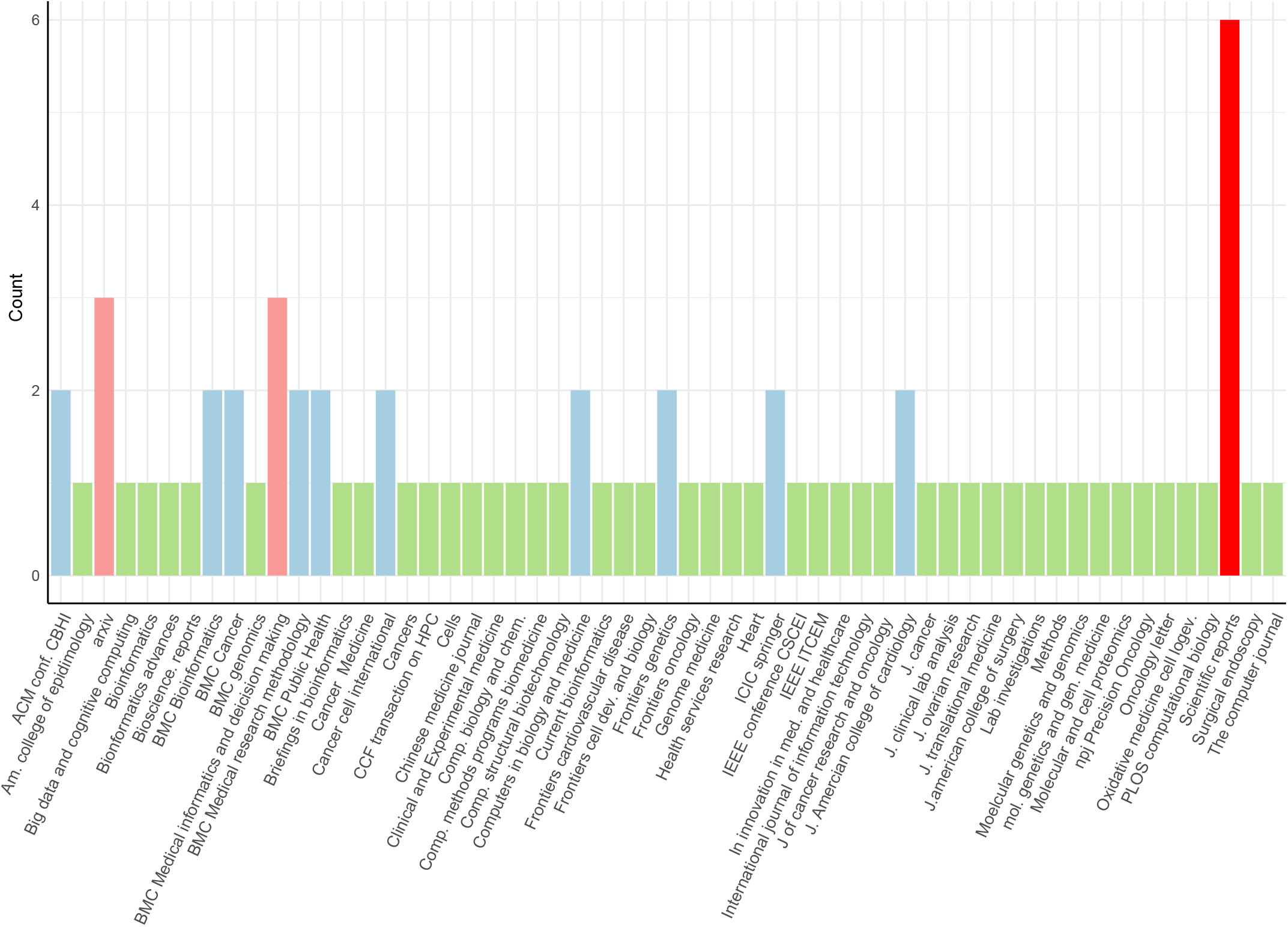
Journal-wise distribution of articles.

**Figure 9.**
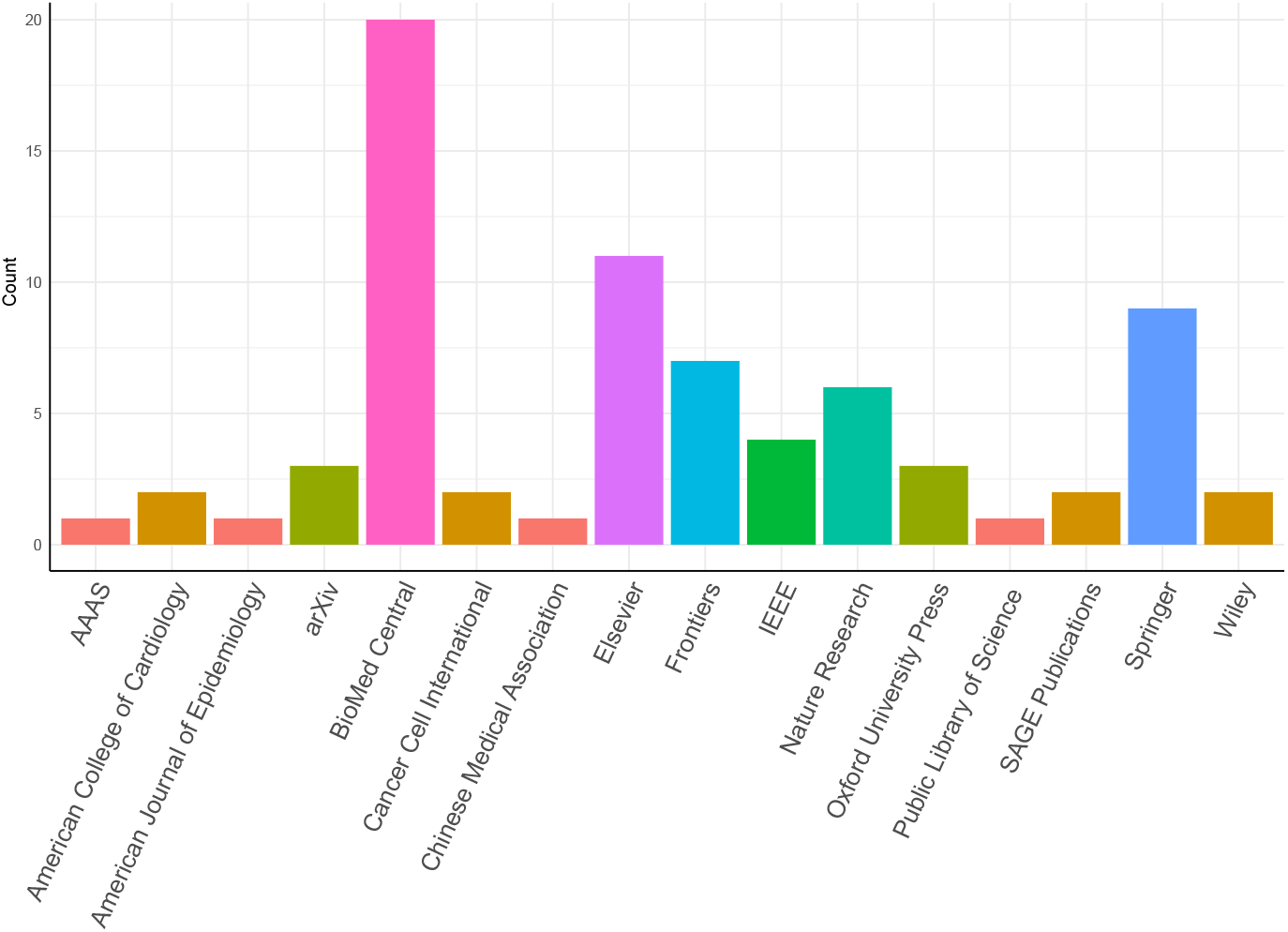
Publisher-wise distribution of articles.

## Discussion

The field of disease survival prediction has become a pivotal aspect of effective healthcare, especially within the domain of precision medicine. Recognizing the significant variability present among patients within specific diseases, there is an increasing demand and development for disease specific survival predictors. Our analysis reveals that researchers place a profound emphasis on predicting survival in cancer as compared to other diseases, and there are compelling reasons behind this focus. First, cancer exhibits significant variability from one patient to another as compared to other diseases, which highlights the imperative need for cancer survival prediction to explore and comprehend the heterogeneity of cancer. Second, cancer is a leading cause of death worldwide, and effective survival prediction can aid in early detection and intervention, potentially saving lives. Third, a huge amount of data sources are developed to make cancer-related data publicly available to accelerate and optimize cancer-related research.

Furthermore, to analyze the trajectory of the disease, researchers place great focus on studying different survival endpoints that suit the respective research setting i.e., treatment, progression, recurrence, and death. Among 4 different survival endpoints i.e., OS, DFS, BC, and PFS, OS is often emphasized more in survival prediction studies. Despite the prime focus on OS, the significance of other survival endpoints in understanding disease trajectories cannot be understated. These survival endpoints help to analyze different characteristics of diseases such as understanding treatment efficacy and durability, treatments that not only extend life but also effectively manage the course of the illness, and markers responsible for disease recurrence. The lack of research in other survival endpoints opens up new research avenues for the AI experts to develop novel methods that can help explore various characteristics related to disease.

Although both public and private databases have been utilized in survival prediction studies, yet the preference for public databases stems from their accessibility and the wealth of information they provide. For instance, TCGA^17^ offers a vast array of genomic and clinical data across different cancer types. This invaluable resource aids researchers in developing accurate survival prediction models. Likewise, GDC^18^ and GEO^130^ offer comprehensive datasets that encompass a wide range of diseases, making them appealing choices for various research endeavors. Furthermore, a crucial observation regarding private data sources is that they are not universally accessible. This argument is supported by the limited accessibility of omics datasets related to cardiovascular diseases. Despite a singular study employing omics data for survival prediction in cardiovascular diseases, the challenge lies in the difficulty of retrieving the original data. Authors often refrain from sharing their datasets, and obtaining access to databases requires extensive proposals, adding a layer of complexity to the development of novel survival prediction pipelines for cardiovascular diseases. This obstacle may impede the advancement of innovative survival prediction pipelines for cardiovascular disease.

Overall, the use of omics and clinical data in survival prediction tools marks a significant stride toward precision medicine. The distribution of omics types in survival prediction studies reveals a preference for mRNA, methylation, microRNA, and CNV across various cancer subtypes. In addition, the limited number of multiomics based survival prediction studies in cardiovascular diseases hinders definitive conclusions on the importance of specific omics types. Disease-specific patterns highlight the importance of tailored clinical markers, prominently seen in cancer studies with a focus on tumor stage and histological type. Treatment-related features, notably chemotherapy and radiotherapy, underscore the impact of therapeutic interventions on survival predictions. Moreover, clinical features along with omics data with diverse molecular aspects are utilized together to improve the performance of survival prediction models. Diverse survival prediction research accentuates the pivotal role of leveraging patient information, such as medical history, demographics, disease-related features, and diagnostic records. This trend reflects an increasing recognition of the potential of clinical data in not only understanding disease progression but also in guiding personalized treatment strategies and enhancing patient care. A recent benchmark study on survival prediction models with multiomics and clinical data also shows the significant role of clinical data in survival prediction across multiple cancer subtypes^45^.

In addition, our analysis reveals that increasing the total number of data modalities does not necessarily offer improved survival predictions, yet data modalities are quite specific to the disease and survival endpoints. Therefore, the selection of data modalities should be made very carefully as rather than improving the overall performance it can induce undesirable noise in the analysis.

One of the common problems in survival analysis is data censoring. Censoring arises when there is incomplete information about the time points and/or events of some subjects in a study. There are different types of censoring i.e. I) Right Censoring is the most common type of data censoring, where an event does not occur for some subjects by the end of study or by the last time point at which data is collected. For example, a subject withdraws from the study or there is a lost follow up for a specific subject II) Left Censoring is the least common type of censoring where the event may occur before the start of the study or during the data collection phase. III) Interval Censoring arises when the event of interest occurs in a time interval but the exact time point is not known. In survival analysis, three assumptions are taken into account to infer censored data i.e., II) Independent Censoring: assumes that the censoring times for multiple subjects are independent of each other. II) Random censoring assumes that the time t at which individuals are censored must be random and the failure rate for subjects who are censored is assumed to be equal to the failure rate for subjects who remained in the risk set who are not censored. III) Non-informative censoring occurs if the distribution of survival times (T) provides no information about the distribution of censorship times (C), and vice versa. Although, data censoring is quite important in terms of survival prediction, yet it has been discussed and dealt with properly in the existing studies. We recommend to incorporate comprehensive details of data censoring in future survival prediction studies. Particularly details on how each type of data censoring is handled should not be neglected.

Our analysis of the utilization of feature engineering methods raises two crucial points. First, even though a plethora of methods have been already tested for various survival prediction studies, autoencoder based methods tend to reduce the dimensionality of omics data modalities more efficiently. In addition, the rest of the methods work much better with clinical features. The success of feature engineering approaches is contingent upon the chosen technique with the inherent properties of the data. This highlights the importance of large-scale benchmark studies in guiding the selection of feature engineering strategies for the development of accurate predictive pipelines.

With an aim to evaluate the performance of predictive pipelines, diverse types of evaluation measures have been developed. Each evaluation measure addresses a specific aspect of survival prediction models, precluding the possibility of any single metric being universally ideal for a comprehensive evaluation of survival prediction. For instance, Cindex estimates the robustness and discriminatory power of the survival prediction model. In addition, BS and IBS measure the accuracy of a model on time distribution. Moreover, log-rank p-value evaluates the potential of the model by testing the differences in different survival groups. Although these measures are the most commonly utilized, there are diverse other evaluation measures for similar purposes i.e., restricted mean survival time (RMST), odds ratio^22^, Kappa for inter-rater reliability^107^, integrated absolute error (IAE), integrated square error (ISE), mean absolute error (MAE), integrated AUC (IAUC) time-dependent integrated discrimination improvement, and time-dependent net reclassification improvement (NRI). Furthermore, while these individual measures provide valuable insights, it is noteworthy to mention that their collective application offers a more comprehensive evaluation. Therefore, we recommend utilizing multiple evaluation measures to assess discrimination and calibration of survival prediction models.

## Methodology

This section explains different steps or stages of preferred reporting items for systematic review and meta-analyses (PRISMA) strategy^161^, which is used to gather relevant papers on survival analysis. Figure 10 provides a visual representation of various stages form PRISMA that are summarised in the following subsections.

**Figure 10.**
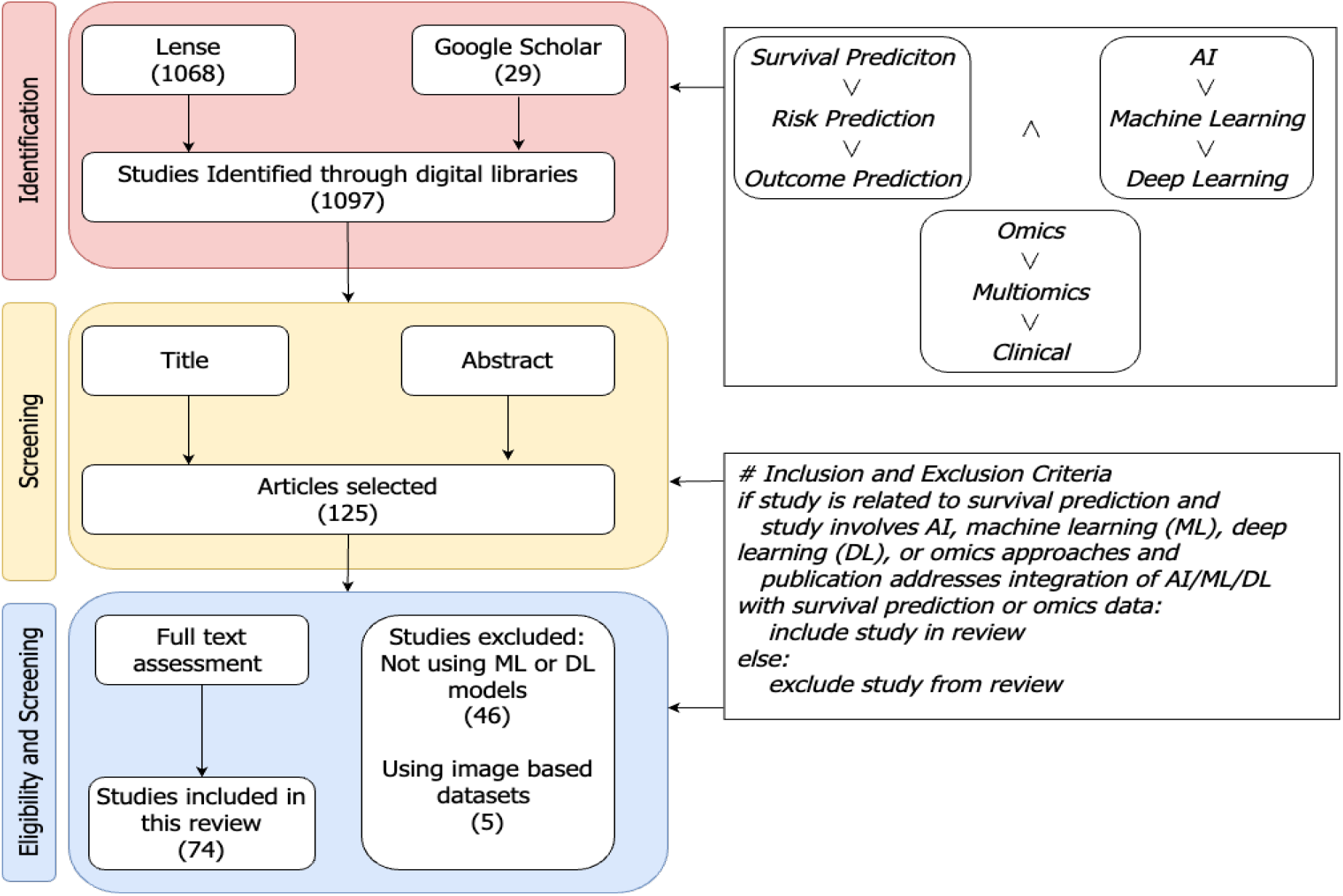
A step-by-step process for articles search and their inclusion or exclusion criteria to generate a set of studies for further in-depth trends analysis

### Search Strategy

In Figure 10, the identification stage illustrates combinations of different keywords that are used to search research articles. The keywords block has two different types of operators ‘∧’ and ‘∨’ operators. On the basis of these operators one keyword from each block is selected and various search queries are formulated such as, *“SURVIVAL PREDICTION AND AI AND OMICS”, “SURVIVAL PREDICTION AND AI AND Multiomics”, “SURVIVAL Machine Learning AND OMICS”*, and so on. These queries are utilized in literature search engines like lens (https://www.lens.org/), and Google Scholar for literature search from Jan 2020 to Jul 2023.

### Screening Strategy

With an aim to retain literature related to survival prediction, two different screenings are performed on the basis of the following criteria;

- Articles that do not make use of only image-based datasets for survival prediction.
- Articles that do not make use of ML, DL, or statistical methods for survival prediction.
- Articles with closed access.

Initially, guided by the title and abstract of the articles, more than 900 studies are discarded. Subsequently, at the final step, based on a comprehensive review of the full text a second screening is performed, resulting in the exclusion of an additional 20 studies. Ultimately, 74 papers are selected for the final comparison and discussion of survival prediction.

## Data Availability

The data is present in different github repositories and public databases

## Additional information

Supplementary tables can be found in additional files. Table S1 entails information about survival prediction studies. In addition, Table S2 presents distribution of data modalities across survival prediction studies. Table S3 shows distribution of survival endpoints across different studies. Table S4 provides a short summary of each study included in this review paper. Table S5, S6, and S7, present information about evaluation measures, journal and publisher wise distribution of survival prediction literature.

## Author contributions statement

A.A. and M.N.A. conducted the literature review, V.S., D.A., and A.S. analyzed the results. All authors reviewed the manuscript.

## Competing interests

The authors declare no competing interests.

## Notes

### Competing Interest Statement

The authors have declared no competing interest.

### Funding Statement

This study did not receive any funding

